# Optimizing Lassa Fever Management using a community pathogen load modeling approach

**DOI:** 10.64898/2025.12.17.25342529

**Authors:** Praise-God Madueme, Faraimunashe Chirove

## Abstract

Lassa Fever control remains a daunting task for authorities in poorly resourced settings where the costs of implementing the control strategies remain high as the disease has multiple hosts and environmental spread. An important metric based on community pathogen load may be useful in estimating the level of control over all in the community in order to budget for the costs of control effectively. We developed a model that accounts for the community contribution of Lassa viral load in humans, rodents as well as the environment accounting for Community Pathogen Load incorporating three control strategies. The model was calibrated and fitted to the Nigerian data and optimized to establish the most cost-effective strategy using cost-effective analysis. Our results suggested that targeting the human community pathogen load remains an important control focus but the control of rodent contribution was equally important. Overall, the combination of three control strategies was the best control measure that is cost-effective for curbing Lassa fever in the population.

## 1 Introduction

Lassa fever continues to pose a serious threat to life in sub-Saharan Africa, where outbreaks occur and are strongly linked to poverty and poor sanitation. Over the last decade, the number of suspected and confirmed cases of Lassa fever infection has increased. Statistics from World Health Organization (WHO) and Centre for Disease Control and Prevention (CDC) reports show an estimate of 100,000 to 300,000 Lassa virus infections in West Africa annually. A good example of this is that cases of Lassa fever in Nigeria are on the rise from 430 suspicious cases in 2015 to 900, 700, 1081, and 5380 suspicious cases in 2016, 2017, 2018, and 2022 respectively [1–5]. The Lassa fever virus epidemic is also increasing in non-endemic areas and countries of the world by traveling abroad. This indicates the urgent need for a better approach to educating healthcare providers on how to detect, diagnose, report, and treat Lassa fever disease and control the spread and clinical manifestations of Lassa virus [6].

Some research has been done on the control of the Lassa virus. Ribavirin is the only recognized antiviral treatment that is used to treat Lassa fever in the early stages of viral infection even though it is not 100% effective [7, 8]. There are other several intervention strategies such as quarantine, early treatment of infected individuals, external protection, fumigation with pesticides, use of condoms to prevent human-to-human transmission during sexual activity, reduction of *Mastomys* rats, and proper sterilization of hospital facilities which have been considered [9–13]. These control strategies have a significant impact on the various transmission routes of the Lassa virus. Even if these strategies are available, the question of funding them can be crucial, especially in developing countries with poor resources and low status where the disease is endemic. Optimal control theory can offer perception into the techniques for controlling the diffusion of Lassa fever at minimum cost [14, 15].

A variety of approaches have been used to model the transmission dynamics and control of Lassa fever [12, 13, 16–19]. In recent times, multiscale models have been used to describe infectious disease systems in terms of the complete pathogen life cycle which represents multiple targets for control. This is because in most zoonotic diseases, the infectious agent (bacteria, virus, and so on) has a complicated life cycle. There are some diseases where part of the life cycle of the pathogen is external to the hosts involved in the transmission of the infection. The disease host is infected by the pathogen’s life stage in the environment. Some other diseases with no free-living pathogen life stage in the environment cause infection to the hosts (vertebrate and vector hosts). Here, the pathogen life cycle is internal to the hosts that are involved in the transmission of the infection [20, 21]. The coupled multiscale models are effective for developing models of infectious disease systems. They are used to identify the processes at the various scales of infectious diseases and how these scales interact. To understand the transmission of infectious disease systems, we require a deep knowledge of these scales namely the between-host scale (associated with disease transmission within the host population where epidemiological processes take place) and the within-host scale (associated with disease dynamics within a single-host where immunological processes take place) [22–25].

Our approach for modeling is similar to the multiscale modeling method used in [20, 21] by introducing the concept of community pathogen load (CPL). Here, CPL refers to the collection of all individual pathogen loads of hosts (humans, animals, vectors, etc.) infected with a particular pathogen (virus, bacteria, fungi, etc.). Lassa fever infection has free-living pathogens in the environment [5] and thus the environment is an important driver. Even though the pathogens do not survive in the environment for a long time, they are continuously replenished by infectious hosts that shed the pathogen in the environment. Since the transmission dynamics of Lassa fever have both direct and indirect routes, it creates a complex system that can be studied using a multiscale modeling approach. Our goal in this work is the introduction of CPL as an additional variable at the between-host scale in modeling the transmission of Lassa fever and incorporating the impact of control strategies on the system. Unlike in [20, 21], our model does not account for all within host stages explicitly but implicitly accounts for the contribution of final infectious within host stages which produces the virus and makes the host capable of transmitting infection to the healthy hosts. At these stages, one can account for the CPL in a community of hosts which is then used as a metric of infection transmission as well as community contribution towards the infection. The remaining part of this work will be presented as follows: Section 2 will be the formulation of the model without control, section 3, a mathematical analysis, section 4, a formulation and analysis of the control model and analysis, section 5, numerical simulations and our discussion of results will be done in Section 6.

## 2 The basic Lassa Fever model with community pathogen load

The total human population is *N*_*H*_ (*t*) and is given as

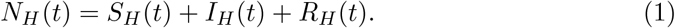

Hsere, *S*_*H*_ (*t*) is the class of susceptible humans, *I*_*H*_ (*t*) refers to the population of all infected humans, and *R*_*H*_ (*t*) is the class of recovered humans. At any time *t*, the constant recruitment of susceptible humans is given as *π*_1_. All humans die naturally at a rate *µ*_1_ and infected humans die due to the Lassa virus at the rate *δ*. Infected humans recover at the rate *ψ*. Susceptible humans contract Lassa fever through the force of infection

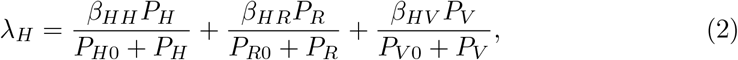

where *β*_*HH*_ is the maximum contact rate of susceptible humans with infected humans, *β*_*HR*_ is the maximum contact rate of susceptible humans with infected rodents, *β*_*HV*_ is the maximum contact rate of susceptible humans with the contaminated environment; *P*_*H*_ is the human community pathogen load (HCPL), *P*_*R*_ is the rodent community pathogen load (RCPL), *P*_*V*_ is the community pathogen load of the virus in the environment (ECPL) and *P*_*H*0_, *P*_*R*0_ and *P*_*V* 0_ are the half-saturation constants of the human, rodent and virus functional response. The HCPL is described as the collection of all individual pathogen loads of humans infected with the Lassa virus in a particular community at a particular time. If one infected human produces *V*_*H*_ virus at the rate *τ*_*H*_, the number of viruses produced by all infected humans is given by *τ*_*H*_*V*_*H*_*I*_*H*_ . Here, *V*_*H*_ is the average population of the Lassa virus within a single infected human host. This total infectious reservoir of humans in the community is our *P*_*H*_. The RCPL is defined as the collection of all individual pathogen loads of rodents infected with the Lassa virus in a particular community at a particular time. If a single infected rodent generates *V*_*R*_ virus at the rate *τ*_*R*_, then *τ*_*R*_*V*_*R*_*I*_*R*_ measures the quantity of Lassa virus generated by all infected rodents. Here, *V*_*R*_ is the average population of the Lassa virus within a single infected rodent host. This total infectious reservoir of rodents in the community is given as *P*_*R*_. The ECPL is the pool that quantifies the total concentration of Lassa virus in the environment at a particular time which is given as *P*_*V*_ .

The total rodent population is *N*_*R*_(*t*) and given as

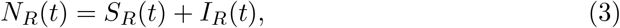

where *S*_*R*_ is the population of rodents susceptible to the virus and *I*_*R*_(*t*) is the population of infected rodents. New recruits move to the rodent population at a constant rate *π*_2_. All rodents die naturally at the rate *µ*_2_ and at a rate *ρ* due to consumption by humans as food. Similarly, susceptible rodents contract the Lassa virus through a force of infection

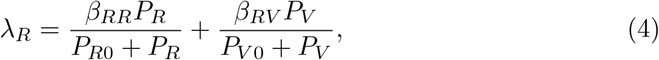

where *β*_*RR*_ is the maximum contact rate of susceptible rodents with infected rodents and *β*_*RV*_ is the maximum contact rate of susceptible rodents with contaminated environment. The rate at which Lassa viral load grows in infected humans is *α*_*H*_ while *α*_*R*_ is the growth rate of Lassa viral load in infected rodents. The classes of infected humans and infected rodents shed the virus in the environment at the rates (1 − *α*_*H*_ ) and (1 − *α*_*R*_) respectively. The natural death/decay rates of the average Lassa viral load within the infected human host, infected rodents, and contaminated environment are *µ*_*P H*_, *µ*_*P R*_, and *µ*_*P V*_ respectively.

The model diagram 1 and the assumptions yields the system of equations:

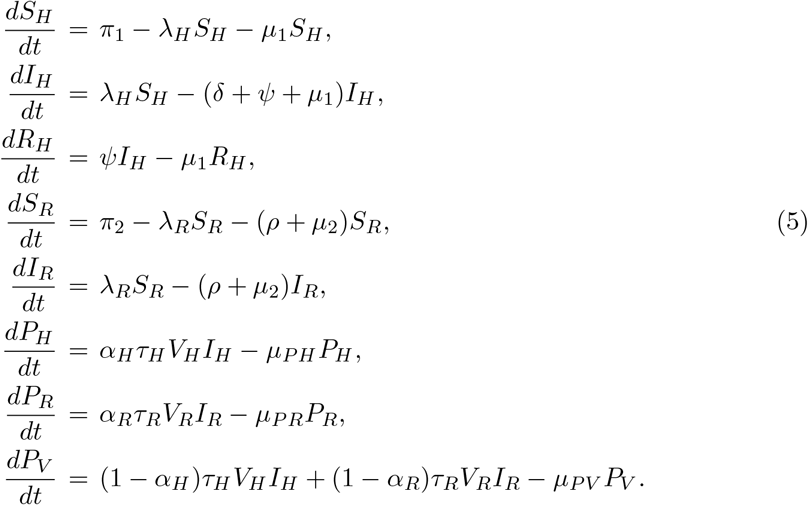

which is subject to the following initial conditions:

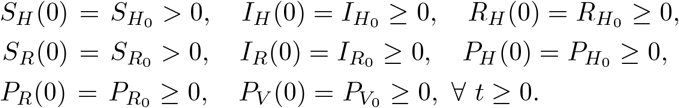

## 3 Model Analysis

### 3.1 Feasible Region

We assume that all parameters for model (5) are non-negative and show that all solutions with non-negative initial conditions remain bounded and non-negative.

#### Theorem 1.

*The solutions S*_*H*_ (*t*), *I*_*H*_ (*t*), *R*_*H*_ (*t*), *S*_*R*_(*t*), *I*_*R*_(*t*), *P*_*H*_ (*t*), *P*_*R*_(*t*), *P*_*V*_ (*t*) *of the system* (5) *are non-negative for t* ≥ 0 *in* Ω *and the region*

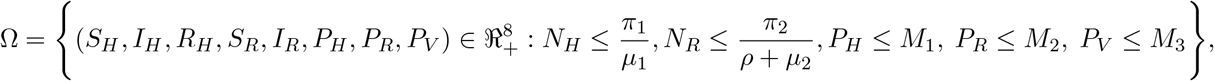

*is positively invariant and attracting*.

*Proof*. We consider the first equation of (5),

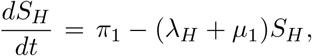

which is solved to obtain

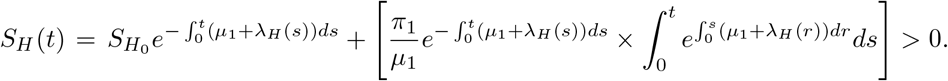

Also, for the solution component of *I*_*H*_ (*t*), we suppose that there exists a first time *t*_1_ such that 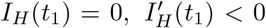 and the rest of the variables are non-negative for 0 *< t*_1_ *< t*. The second equation of system (5) gives

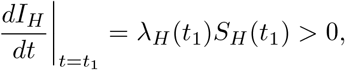

which is a contradiction, so *I*_*H*_ (*t*) ≥ 0, ∀ *t* ≥ 0. Using a similar approach, it is easy to show that *R*_*H*_ (*t*), *S*_*R*_(*t*), *I*_*R*_(*t*), *P*_*H*_ (*t*), *P*_*R*_(*t*), *P*_*V*_ (*t*) are non-negative. Hence, all solutions of (5) are non-negative.

Also, adding the first three equations in the system (5) gives

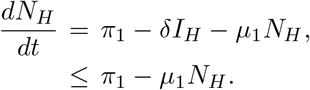

Then

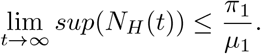

Adding the equations four and five in the system (5) gives

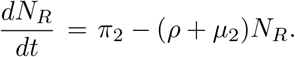

Then

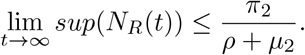

Using a similar technique, we can show that the remaining model variables are non-negative. So, all feasible solutions of our model (5) are non-negative and enter the invariant attracting region

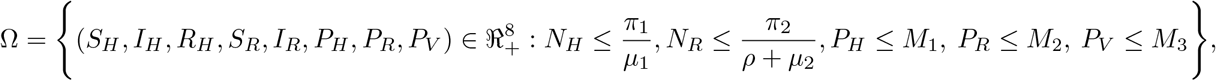

where

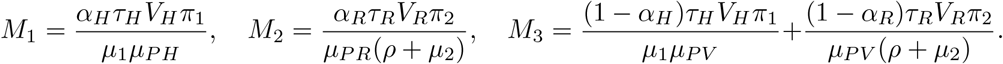

Ω is positively invariant and attracting and this guarantees that existence, uniqueness, and continuation results for model (5) hold in this region and all solutions which start in Ω also stay inside it for *t* ≥ 0. Our model (5), is therefore mathematically and epidemiologically meaningful. □

### 3.2 Equilibrium and Stability Analysis

We obtain the equilibrium states of the model by setting the right-hand side of model (5) to zero. At the disease-free equilibrium, there is no Lassa virus and hence no infection in the human and rodent population. The disease-free equilibrium of system (5) is given by

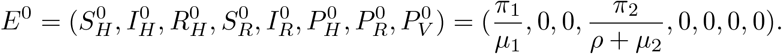

We obtain the basic reproduction number, *R*_0_ of model (5) using the next generation operator approach [26]. Model (5) can be written in the form

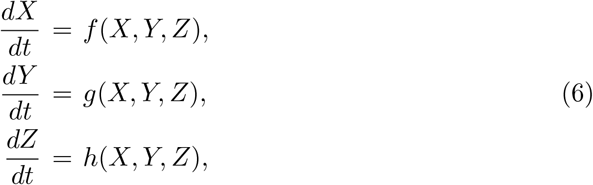

where

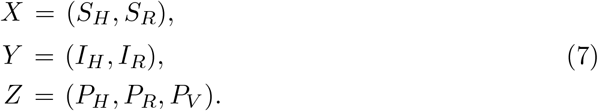

The components of *X* represent the number of susceptibles, the components of *Y* signify the number of infected individuals that do not spread the virus while the components of *Z* signify the number of individuals that can spread the virus. We define

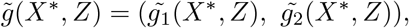

with

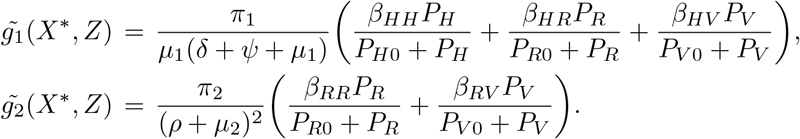

Let 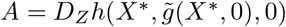 and further assume that *A* can be written as *A* = *M* −*D*, where *M* ≥ 0 and *D* ≥ 0 a diagonal matrix. Then *A* will be

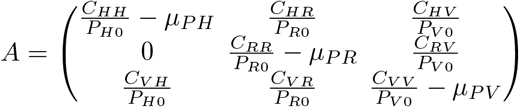

where

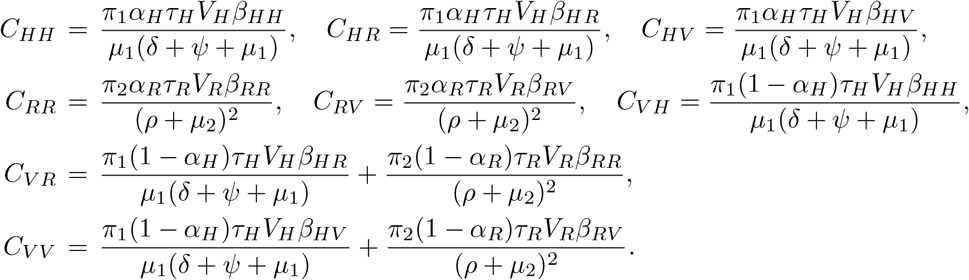

Because *A* = *M* − *D*, we infer that matrices *M* and *D* to be

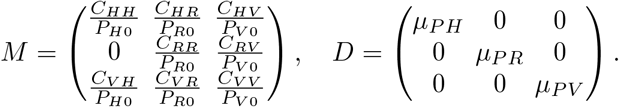

The basic reproduction number is the spectral radius (dominant eigenvalue) of the matrix *MD*^−1^. So

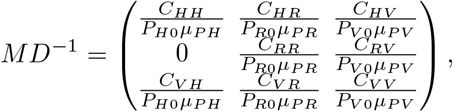

and

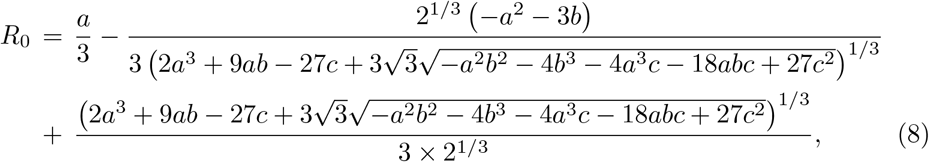

where

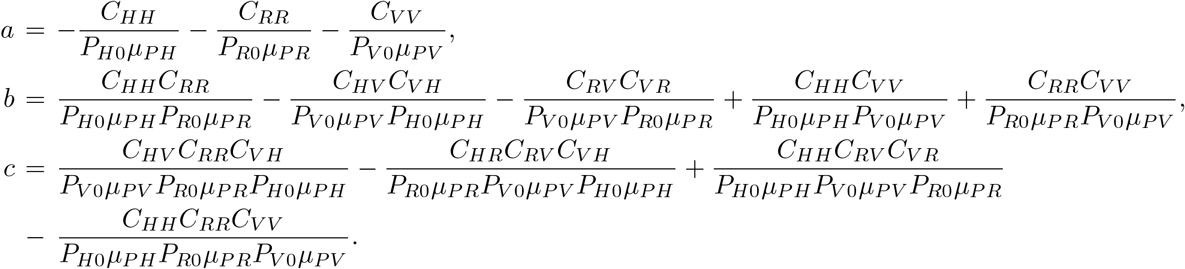

#### Theorem 2.

*The disease free equilibrium, E*^0^, *for model* (5) *is globally asymptotically stable if R*_0_ ≤ 1.

*Proof*. We apply the approach by Kamgang and Sallet [27] to investigate the global stability of the disease-free equilibrium. It suffices to show that model (5) satisfies hypotheses **H1** to **H5** of the global stability theorem. Let **x**_**1**_ denote the density of the non-infected classes of the model and **x**_**2**_ be the density of the infected classes; that is, **x**_**1**_ = (*S*_*H*_, *R*_*H*_, *S*_*R*_) and **x**_**2**_ = (*I*_*H*_, *I*_*R*_, *P*_*H*_, *P*_*R*_, *P*_*V*_ ). The domain Ω is a compact set and obviously positively invariant.

We consider the sub-system 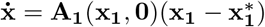:

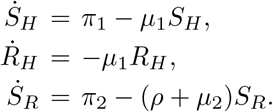

This is a linear system which is globally asymptotically stable at *E*^0^, satisfying the hypotheses **H1** and **H2**.

The matrix **A**_**2**_(**x**) is given by

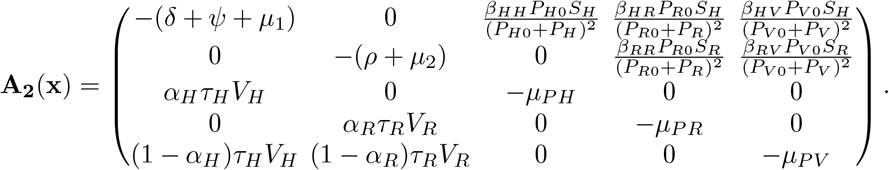

The matrix **A**_**2**_(**x**) is Metzler and irreducible for any **x** ∈ **Ω**, which satisfies hypothesis **H3**.

For hypothesis **H4**, there is a maximum matrix which is uniquely realized in Ω at *E*^0^. This maximum matrix, 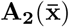, the block of the Jacobian at *E*^0^, is given by

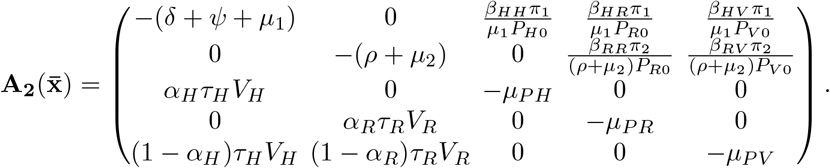

This shows the situation in Corollary 4.4 [27], where the maximum is attained at *E*^0^. For hypothesis **H5**, we require that 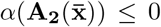 where 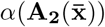 is the stability modulus (spectral bound) of the square matrix 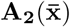. We write 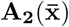 as a block matrix,

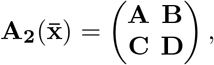

where

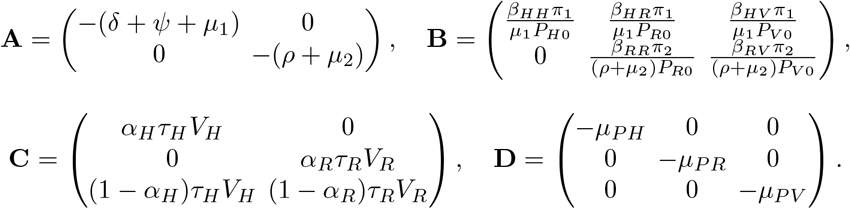

**A** is a Metzler stable matrix and the condition 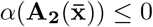 is equivalent to *α*(*D* − *CA*^−1^*B*) ≤ 0. Let

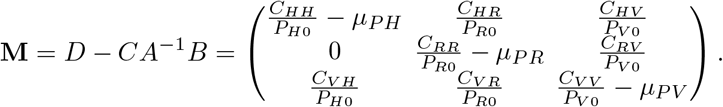

where

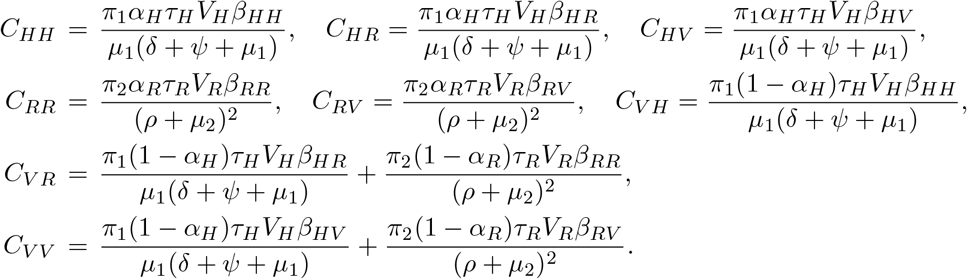

*M* is a stable Metzler matrix if and only if *R*_0_ ≤ 1, that is 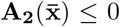 if and only if *R*_0_ ≤ 1. Having satisfied hypothesis **H1** to **H5**, *E*^0^ is globally asymptotically stable if and only if *R*_0_ ≤ 1. □

The endemic equilibrium of system (5) is given by

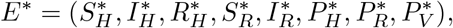

where

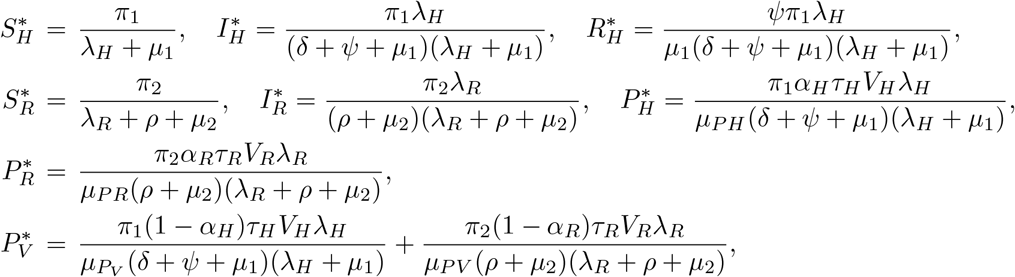

and

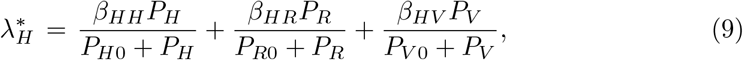

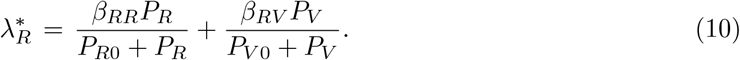

We write equations (9) and (10) as

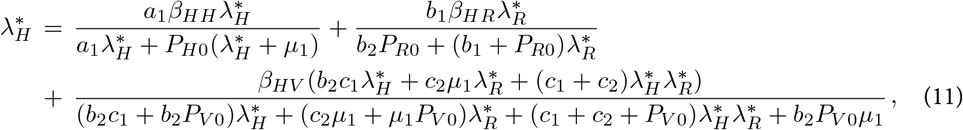

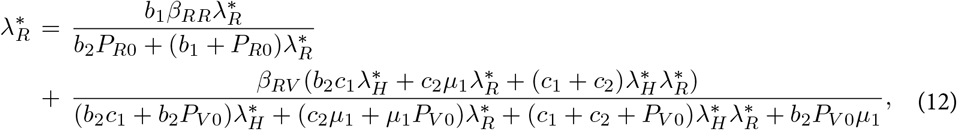

where

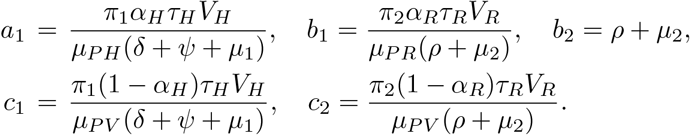

Since the state variables are expressed in terms of 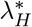 and 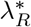, we use the approach in [28, 29] to obtain positive equilibrium points of the model by finding the fixed points of equations (11) and (12) as

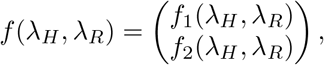

where

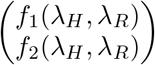

corresponds to the right hand sides of equation (11) and (12).

#### Theorem 3.

*There exists a unique fixed point* 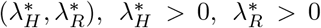 *which satisfies*

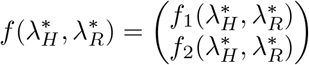

*and corresponds to the endemic equilibrium point E*^*^ *[28, 29]*.

*Proof*. From the first equation, we fix *λ*_*R*_ *>* 0 and look at the real-valued function depending on *λ*_*H*_ :

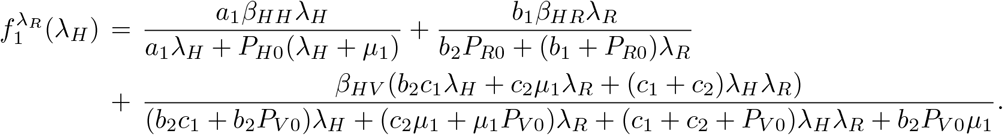

We have that

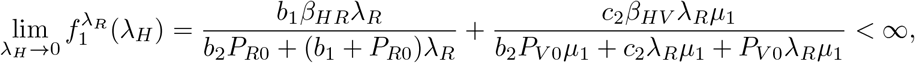

and

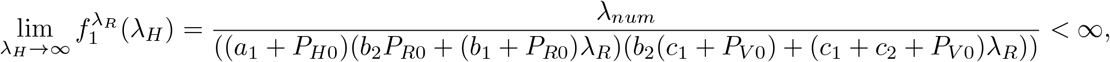

where

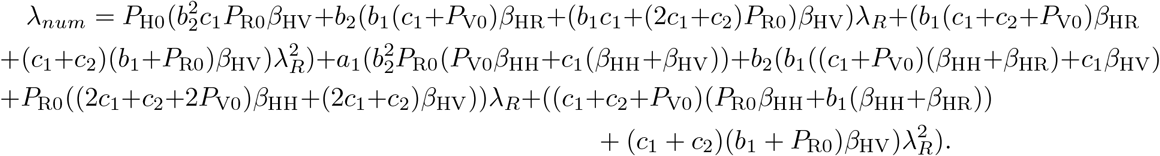

It follows that 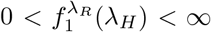 which implies that 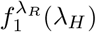 is bounded for every fixed *λ*_*R*_ *>* 0.

Next,

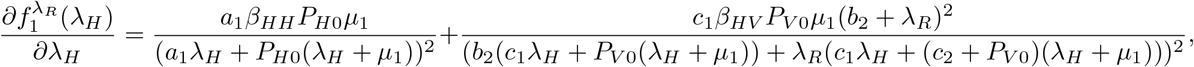

and

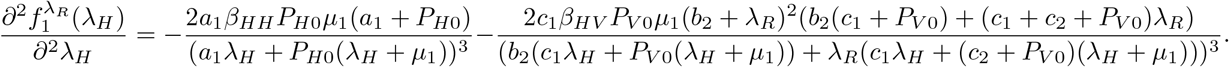

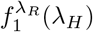 is an increasing concave down function since 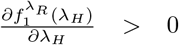 and 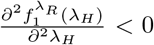. Hence, there is no change in concavity of *f*_1_ in the bounded domain. It follows that there exists a unique 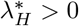 which satisfies 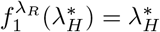.

For this 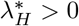, we look at the real-valued function depending on *λ*_*R*_:

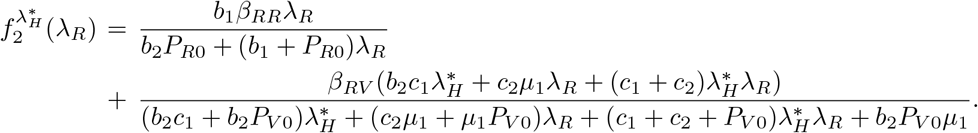

Then,

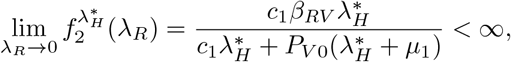

and

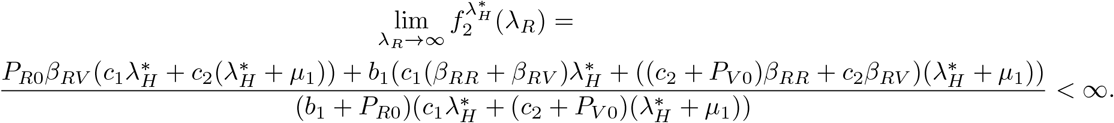

It follows that 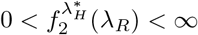 which implies that 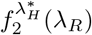 is bounded for every fixed 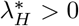.

Next,

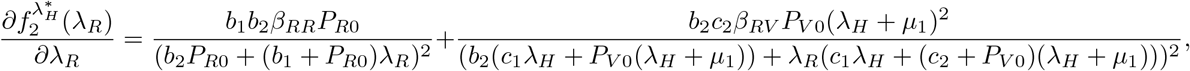

and

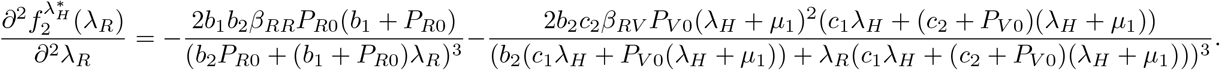

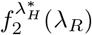 is an increasing concave down function since 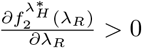 and 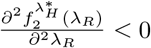. Hence, there is no change in the concavity of *f*_2_ in the positive domain. It follows that there exists a unique 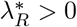 which satisfies 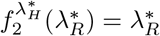.

Therefore, there is a fixed point 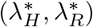 which corresponds to the endemic equilibrium point *E*^*^. □

The stability conditions of *E*^*^ are determined by the eigenvalues of the Jacobian matrix of equations (11) and (12) given as

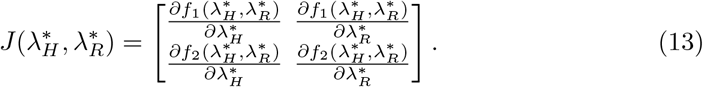

#### Theorem 4.

*[30] Let λ*_*i*_, *i* = *H, R, be the eigenvalues of the Jacobian matrix* 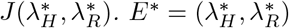 *is asymptotically stable if* |*λ*_*i*_| *<* 1.

*Proof*. Let

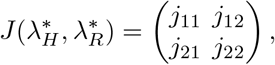

where

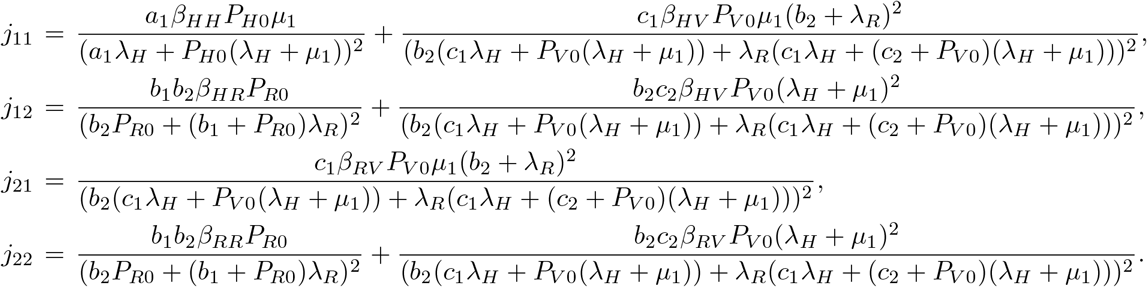

|*λ*_*i*_| *<* 1 corresponds to

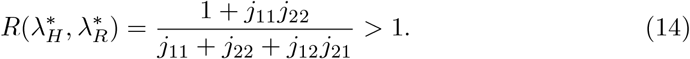

The fixed point 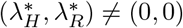 is locally stable when 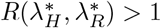. □

## 4 The Control Model

There are numerous ways that Lassa fever is transmitted [31]. The transmission of this virus has been managed by a number of intervention methods. For instance, external shielding can prevent human-to-human transmission through infected medical equipment or contact with bodily fluids (such as the use of personal protective equipment, and condom use). Numerous mathematical models have recently been applied to understand better methods for preventing the spread of infectious diseases, such as isolation, immunization, and fumigation [32, 33]. If there is an effective therapy, the majority of infectious diseases are significantly decreased. Patients with infections have primarily received symptomatic and supportive care; a small number of patients received particular treatment with Lassa-immune plasma, with varying degrees of efficacy. Only when started early in the course of the illness may ribavirin, an antiviral medicine, effectively treat Lassa fever. It is interesting to note that when an infected person gets better, they develop lifelong immunity (they are not susceptible again). It is important to explore ways to stop the spread of Lassa fever using treatment as a control intervention method since effective treatment is a good control strategy [9, 11]. The likelihood that humans will come into touch with diseased rats is increased by a number of circumstances, including poor sanitation and unhealthy human behavior. By implementing better hygiene procedures and sanitizing facilities as a rodent control measure, we can lower this contact rate. A single control technique won’t be sufficient in the event of an outbreak, even while control treatments taken one at a time do have some effect on lowering the population’s rate of secondary infections. To reduce the viral load in the system, it is crucial to integrate a combination of intervention measures [13]. In this work, infection is a result of a community contribution and not as individual aggregates. We consider the impact of time-dependent control measures on the community level of contribution using a combination of multiple intervention strategies (treatment, fumigation, rodent control) on our control-free model and also perform the optimal control analysis. To introduce treatment (using ribavirin) in the basic model (5), we assume that infected humans are treated at the rate *u*_1_, for 0 ≤ *u*_1_(*t*) ≤ 1. We also incorporate the rodent control (using mouse traps) rate *u*_2_, 0 ≤ *u*_2_(*t*) ≤ 1 and the fumigation to control the virus in the environment (using pesticides) rate *u*_3_, 0 ≤ *u*_3_(*t*) ≤ 1. The system of equations for our control model is:

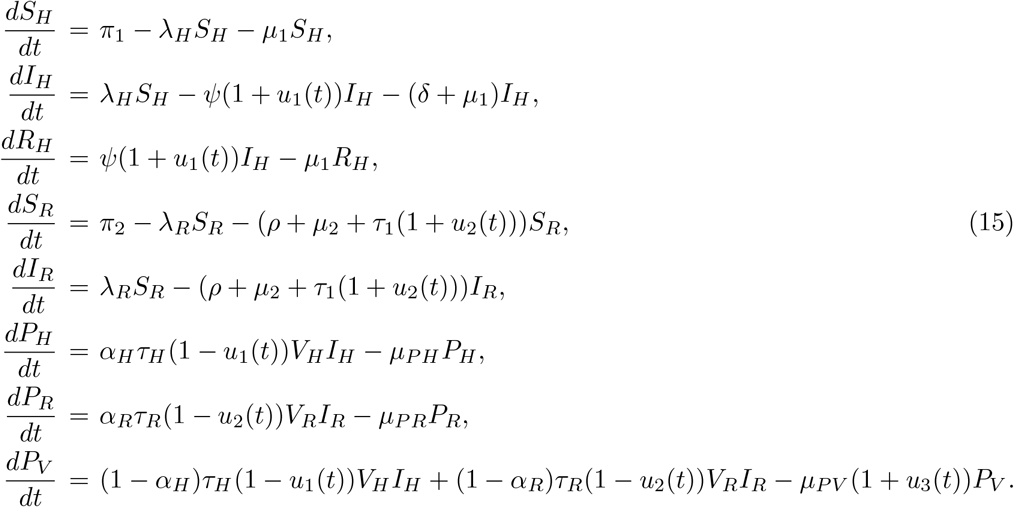

### 4.1 Optimal Control Problem and Analysis

The best control measure for curbing the spread of Lassa fever is the multiple control intervention technique [13]. However, it is obvious that some communities or areas where this disease occurs may be faced with limited resources needed to combat the menace. This is why optimal control is considered a better intervention scheme that will not just eradicate the disease but also do it with minimum cost [14, 15]. We use the optimal control theory to analyze this problem. In this work, we attempt to reduce the cost of implementing multiple control intervention techniques by taking the control parameters *u*_1_, *u*_2_ and *u*_3_ in the control model (15) as measurable functions of time *t* and then formulate an appropriate optimal control functional that minimizes the cost of implementing the controls subject to the model. Our control scheme is said to be optimal if it minimizes the objective functional:

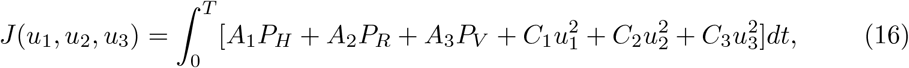

where *A*_1_, *A*_2_, *A*_3_, *C*_1_, *C*_2_, *C*_3_ are balancing coefficients. We use this procedure to minimize the community pathogen load of infected humans, infected rodents, and contaminated environment as well as the cost of applying the controls. The existence of the optimal control triple 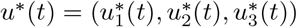 that minimizes our objective functional (16) subject to the state system which is the control model (15) comes from

Fleming and Rishel [34]:

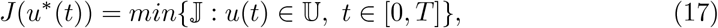

where

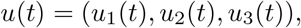

and 𝕌 = {*u*(*t*) : *u*(*t*) are measurable, 0 ≤ *u*(*t*) ≤ 1} is the control set. The Pontryagin’s Maximum Principle [35] introduces adjoint functions that allow us to attach our state system (15) to our objective functional (16). It is used to minimize a Hamiltonian H, by converting the minimization problem of the objective functional state system, with respect to *u*_1_(*t*), *u*_2_(*t*) and *u*_3_(*t*). The Hamiltonian *H* is given by

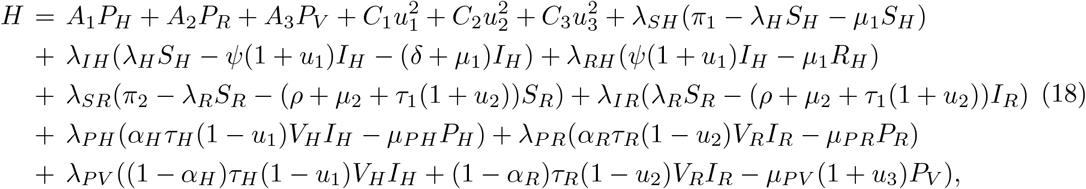

where *λ*_*SH*_, *λ*_*IH*_, *λ*_*RH*_, *λ*_*SR*_, *λ*_*IR*_, *λ*_*P H*_, *λ*_*P R*_, *λ*_*P V*_ are associated adjoints for the states *S*_*H*_, *I*_*H*_, *R*_*H*_, *S*_*R*_,

*I*_*R*_, *P*_*H*_, *P*_*R*_, and *P*_*V*_ respectively. Given the optimal control triple 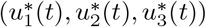 and corresponding states (*S*_*H*_, *I*_*H*_, *R*_*H*_, *S*_*R*_, *I*_*R*_, *P*_*H*_, *P*_*R*_, *P*_*V*_ ), there exist adjoint func-tions satisfying

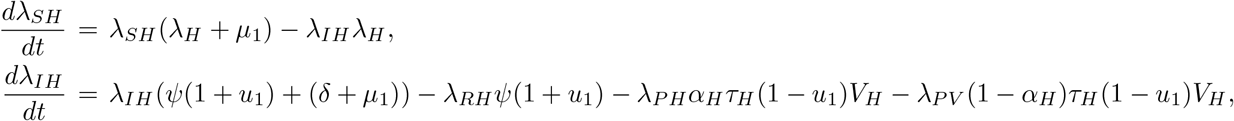

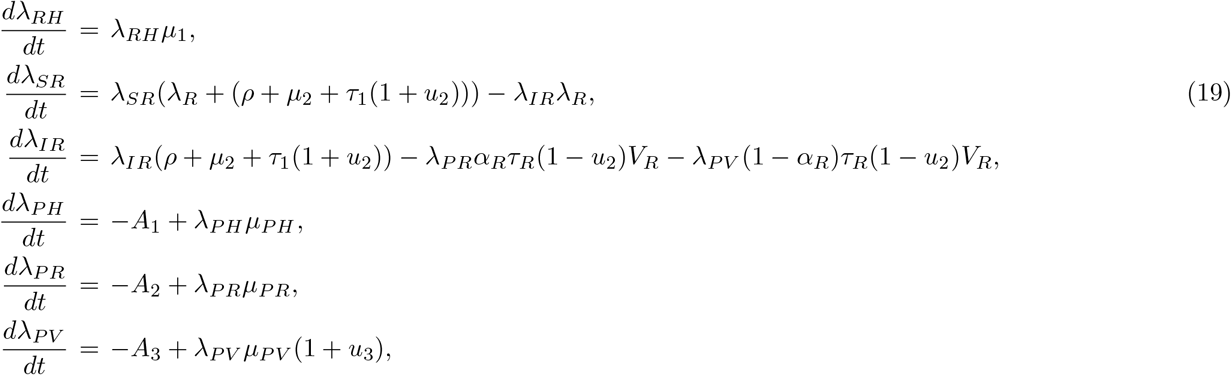

with transversality conditions *λ*_*k*_(*T* ) = 0, for *k* = *S*_*H*_, *I*_*H*_, *R*_*H*_, *S*_*R*_, *I*_*R*_, *P*_*H*_, *P*_*R*_, and *P*_*V*_ .

#### Remark 1.

*The differential equations* (19) *is obtained by:*

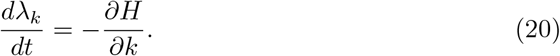

Using the optimality conditions

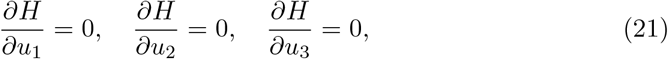

we get

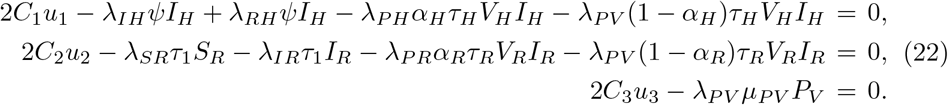

Solving for *u*_1_, *u*_2_, *u*_3_, we have

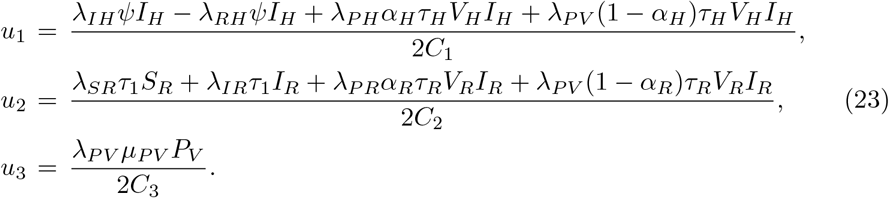

We take the boundary conditions into consideration to get

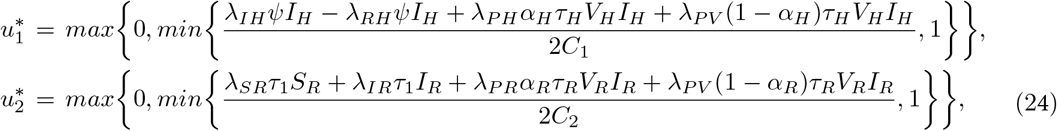

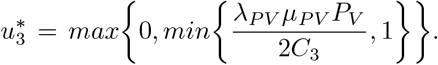

We see that there exists an optimal control triple 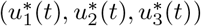 which decreases the spread of Lassa fever disease considering the control intervention strategy with minimum cost. For sufficiently small *t*, the solution to the optimality system of the optimal control problem exists and is unique [36]. Since the state equations have initial conditions and the adjoint equations have final time conditions, we can only solve the optimality system using iterative algorithms and numerical simulations to reveal more details on the trajectory of the optimal control using published data.

## 5 Numerical Simulations

### 5.1 Parameter Estimation

Our focus will be on four states (Ondo, Edo, Ebonyi, Bauchi), Nigeria where the virus has been endemic over the past few years [37]. To estimate our model parameters for the fitting process, we use the available data (weekly recorded cases) on Nigeria’s epidemic site and use the least-squares fitting routine function (scipy optimize) in Python. The method helps to obtain the values of the parameters that are then used in the numerical simulations. The model fit in figure 2 was done using the cumulative cases of our data points and captured using the equation:

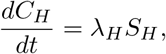

subject to the initial condition *C*_*H*_ (0) = 0.

**Fig. 1:**
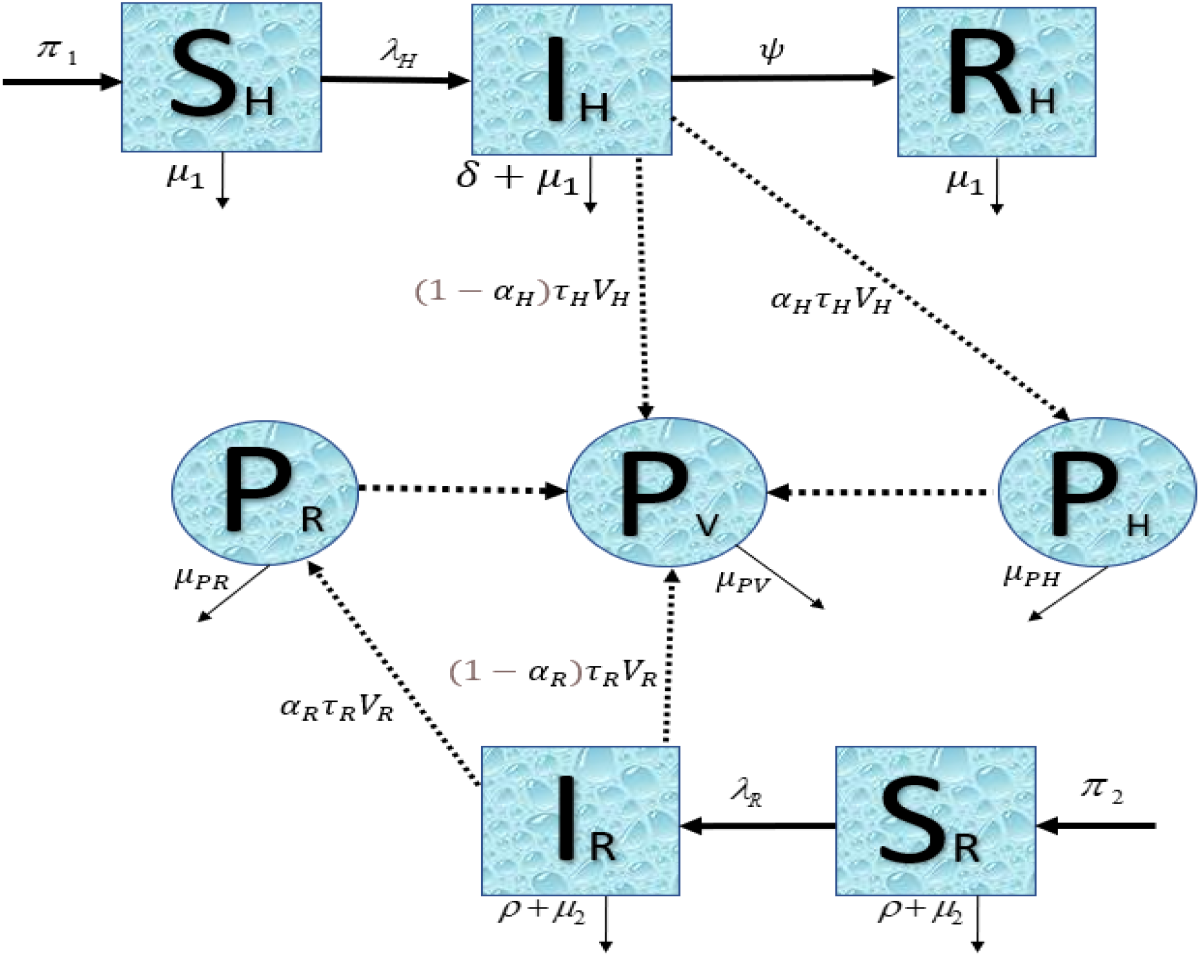
The Lassa Fever infection model diagram with community pathogen load.

**Fig. 2:**
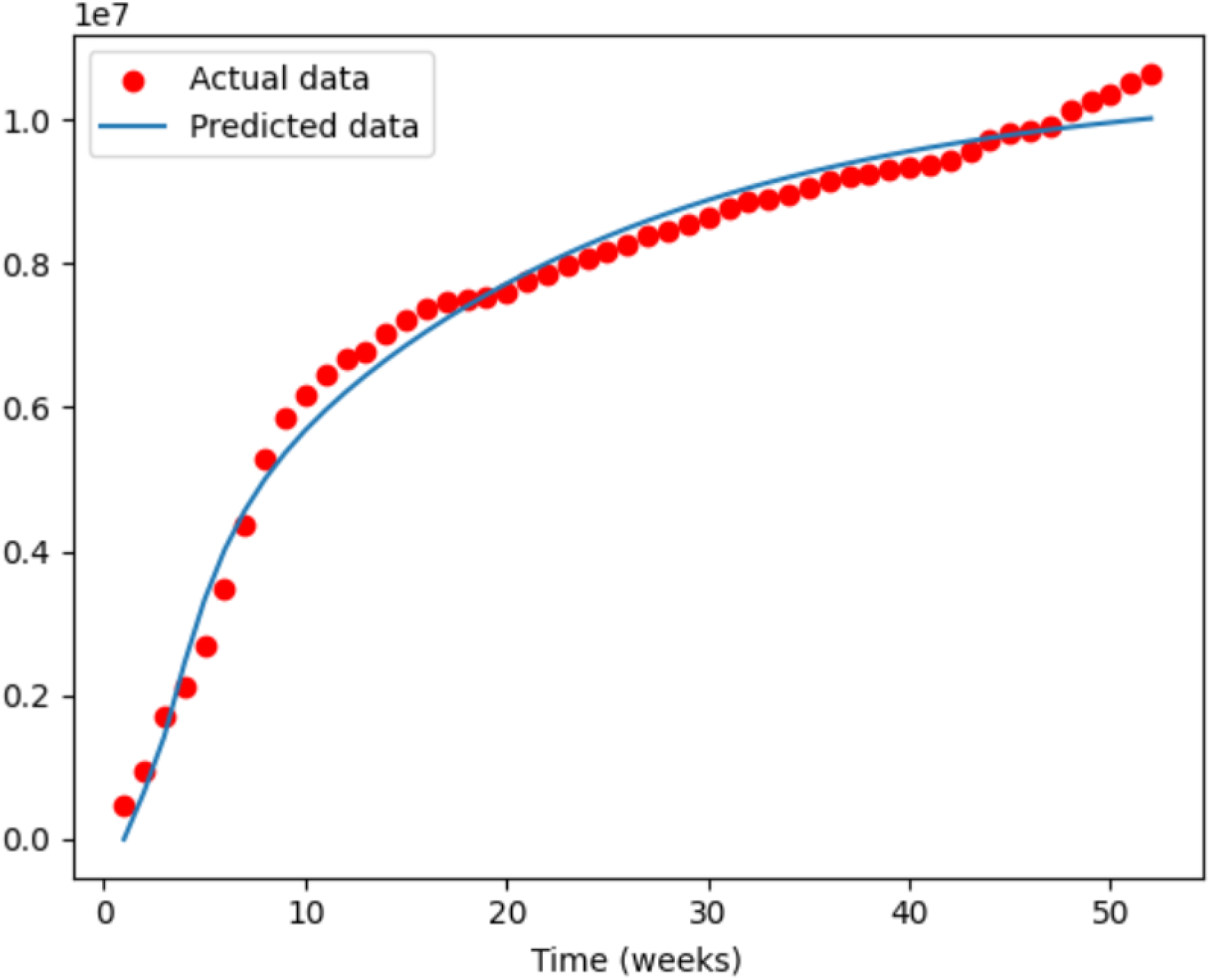
The Cumulative human classes fit real data of the Nigerian states.

In the chosen region, we consider an average population of 19, 978, 662 persons, 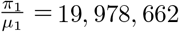. The human natural death rate is 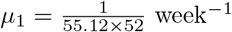 using the average human lifespan in Nigeria as 55.12 [38]. The daily recruitment rate of humans is estimated as *π*_1_ = 19978662 *× µ*_1_ = 6970.37 week^−1^. We consider a hypothetical average population of *Mastomys* rats to be 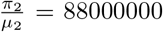, since there is no known quantified estimate of the rodent population. The average lifespan of a rodent is 1 year [39], so we obtain 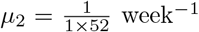. We estimate the rodent recruitment rate to be *π*_2_ = 88000000 *× µ*_2_ = 1692240 week^−1^. For our simulation, we use the following initial conditions: *S*_*H*_ (0) = 19978662, *I*_*H*_ (0) = 48, *R*_*H*_ (0) = 10, *S*_*R*_(0) = 88000000, *I*_*R*_(0) = 100, *P*_*H*_ (0) = 24000, *P*_*R*_(0) = 5000, *P*_*V*_ (0) = 100.

Figure 2 shows that model (15) fits well to the Lassa fever new weekly cases of Nigeria from the 3rd of January to the 28th of December 2022, with data obtained from [1]. Our *R*^2^ = 97% value shows how well the data fits the regression model or the goodness of fit. This value suggests that the model and parameter estimates can be used to explain the underlying trends in the data.

The values of the coefficients in table 2 determine how much emphasis is placed on each term or component in the cost function. They represent the trade-offs and priorities of the optimization problem, allowing you to balance different aspects of system performance according to real-world objectives and constraints. The presence of constraints allows our objective function to penalize deviations from desired trajectories.

**Table 1:** Parameter values and references.

| Parameters | Meaning | Value | Reference |
| --- | --- | --- | --- |
| $\pi_1$ | Human recruitment rate | 6970.37 | Calculated |
| $\pi_2$ | Rodent recruitment rate | 1692240 | Calculated |
| $\mu_1$ | Human natural death rate | 0.0003488891 | [38] |
| $\mu_2$ | Rodent natural death rate | 0.01923 | [39] |
| $\psi$ | Human recovery rate | 0.0000100436708 | Fitted |
| $\beta_{HH}$ | Contact rate of $S_H$ with $I_H$ | 0.0209 | Fitted |
| $\beta_{HR}$ | Contact rate of $S_H$ with $I_R$ | 0.0122 | Fitted |
| $\beta_{HV}$ | Contact rate of $S_H$ with $P_V$ | 0.0121 | Fitted |
| $\beta_{RR}$ | Contact rate of $S_R$ with $I_R$ | 0.3086 | Fitted |
| $\beta_{RV}$ | Contact rate of $S_R$ with $P_V$ | 0.008 | Fitted |
| $\rho$ | Rodent consumption rate | 0.000085714 | [40, 41] |
| $\delta$ | Human disease-induced death rate | 0.00714285 | [37] |
| $P_{HO}$ | $P_H$ when $\beta_{HH}$ is half (half saturation constant) | 8024.574 | Fitted |
| $P_{RO}$ | $P_R$ when $\beta_{HR}$ is half (half saturation constant) | 5000 | Fitted |
| $P_{VO}$ | $P_V$ when $\beta_{HV}$ is half (half saturation constant) | 4975.426 | Fitted |
| $\alpha_H$ | Growth rate of Lassa viral load in infected humans | 0.5158 | Fitted |
| $\alpha_R$ | Growth rate of Lassa viral load in infected rodents | 0.3086 | Fitted |
| $\tau_H$ | Production rate of $V_H$ virus in one infected human | 0.3086 | Fitted |
| $\tau_R$ | Production rate of $V_R$ virus in one infected rodent | 0.1012 | Fitted |
| $V_H$ | Average LF load within a single infected human host | 502.4574 | Fitted |
| $V_R$ | Average LF load within a single infected rodent host | 50.2457 | Fitted |
| $\mu_{PH}$ | Decay rate of LF load within infected human host | 0.0141 | Fitted |
| $\mu_{PR}$ | Decay rate of LF load within infected rodent host | 0.01899 | Fitted |
| $\mu_{PV}$ | Decay rate of LF load in contaminated environment | 0.008977 | Fitted |
| $\tau_1$ | Trapping rate of rodents | 0.000001 | Fitted |
| $u_1$ | Human treatment rate | 0.6914 | Fitted |
| $u_2$ | Rodent control rate | 0.4939 | Fitted |
| $u_3$ | Environment Fumigation rate | 0.3951 | Fitted |

**Table 2:** Costs associated with optimal control variables used for simulations.

| Parameter | Meaning | Value | Reference |
| --- | --- | --- | --- |
| $A_1$ | Weight associated with reducing the HCPL | 1 | Calculated |
| $A_2$ | Weight associated with reducing the RCPL | 3 | Calculated |
| $A_3$ | Weight associated with reducing the VCPL | 1 | Estimated |
| $C_1$ | Weight on the cost of the treatment | $10^3$ | Estimated |
| $C_2$ | Weight on the cost of the rodent control | $10^2$ | Estimated |
| $C_3$ | Weight on the cost of the fumigation | $10^3$ | Estimated |

### 5.2 Simulation Results

In this section, we present the results of our simulation. We show the baseline graphs for all the classes (see figure 3) and the changes in HCPL, RCPL and VCPL without control interventions and with constant controls (see figure 4). We showed the effect of only constant human treatment (*u*_1_ ≠ 0, *u*_2_ = 0, *u*_3_ = 0) on HCPL, RCPL and VCPL (see figure 5), the effect of only constant rodent control (*u*_1_ = 0, *u*_2_ ≠ 0, *u*_3_ = 0) on HCPL, RCPL and VCPL (see figure 6), and the effect of only constant on fumigation (*u*_1_ = 0, *u*_2_ = 0, *u*_3_ ≠ 0) on HCPL, RCPL and VCPL (see figure 7). We also show the numerical simulations of different time-varying control strategies on the HCPL, RCPL, and VCPL (see figures 8 to 14).

**Fig. 3:**
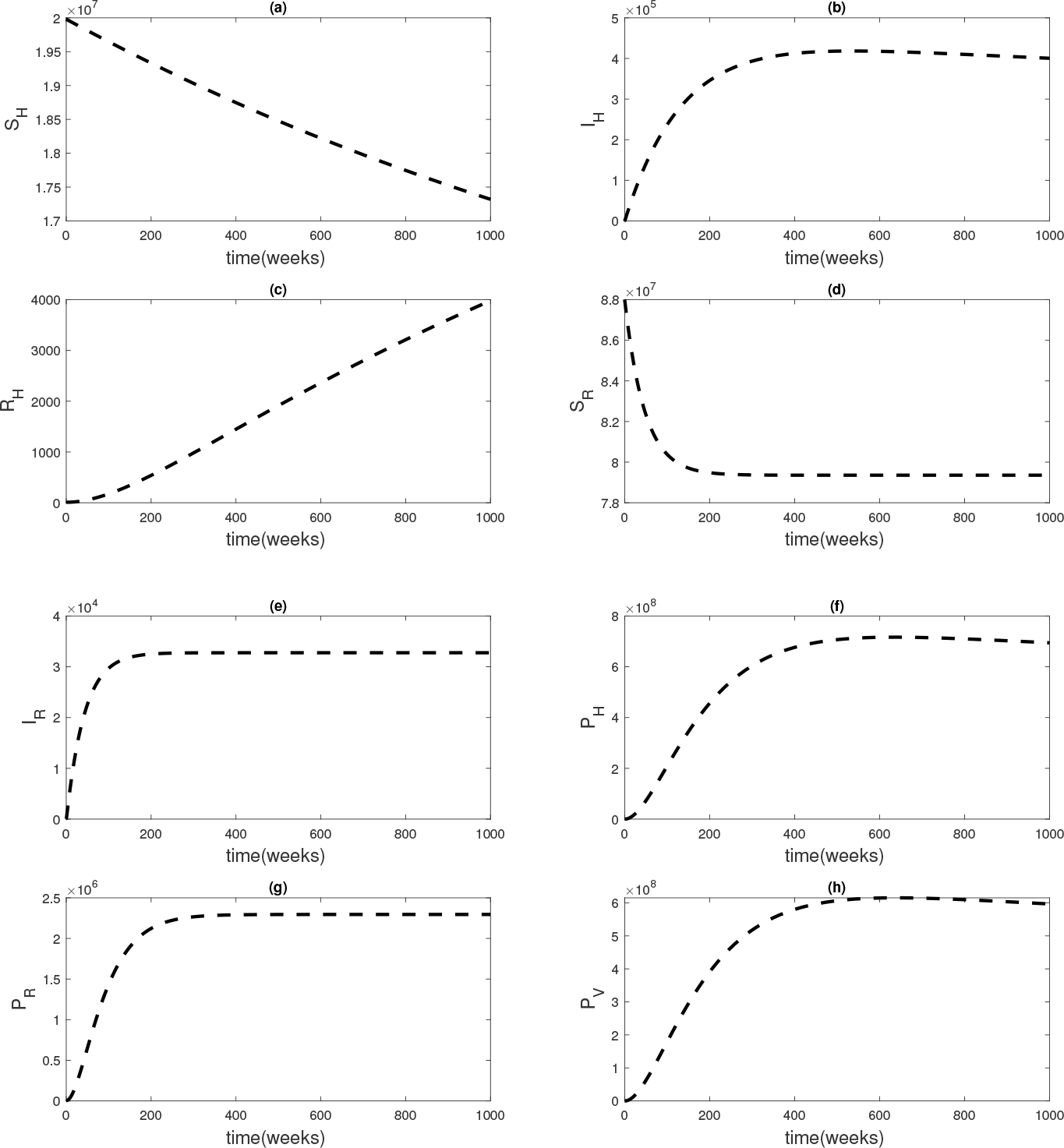
Graphical illustration of model (5) for *R*_0_ greater than unity: *R*_0_ = 3.1868.

**Fig. 4:**
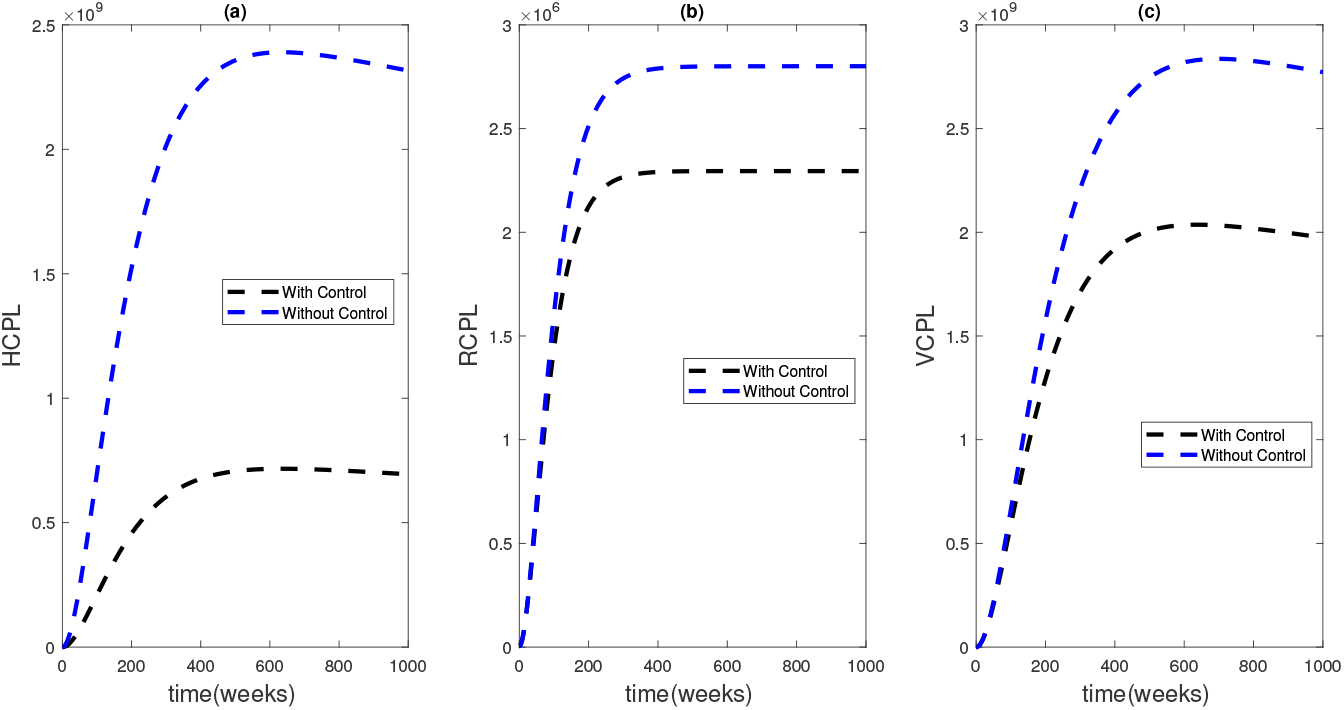
Graphical illustration of the Community Pathogen Load with and without constant control.

**Fig. 5:**
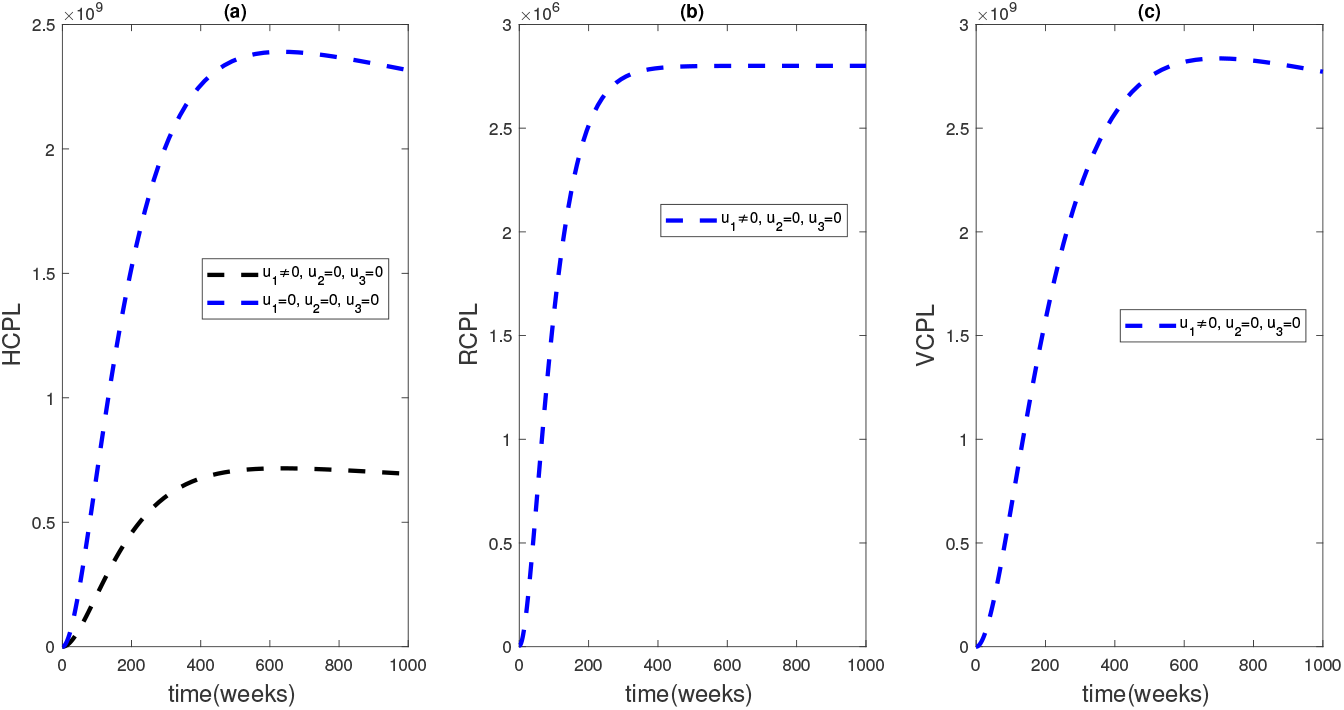
Graphical illustration of the Community Pathogen Load with constant human treatment control.

**Fig. 6:**
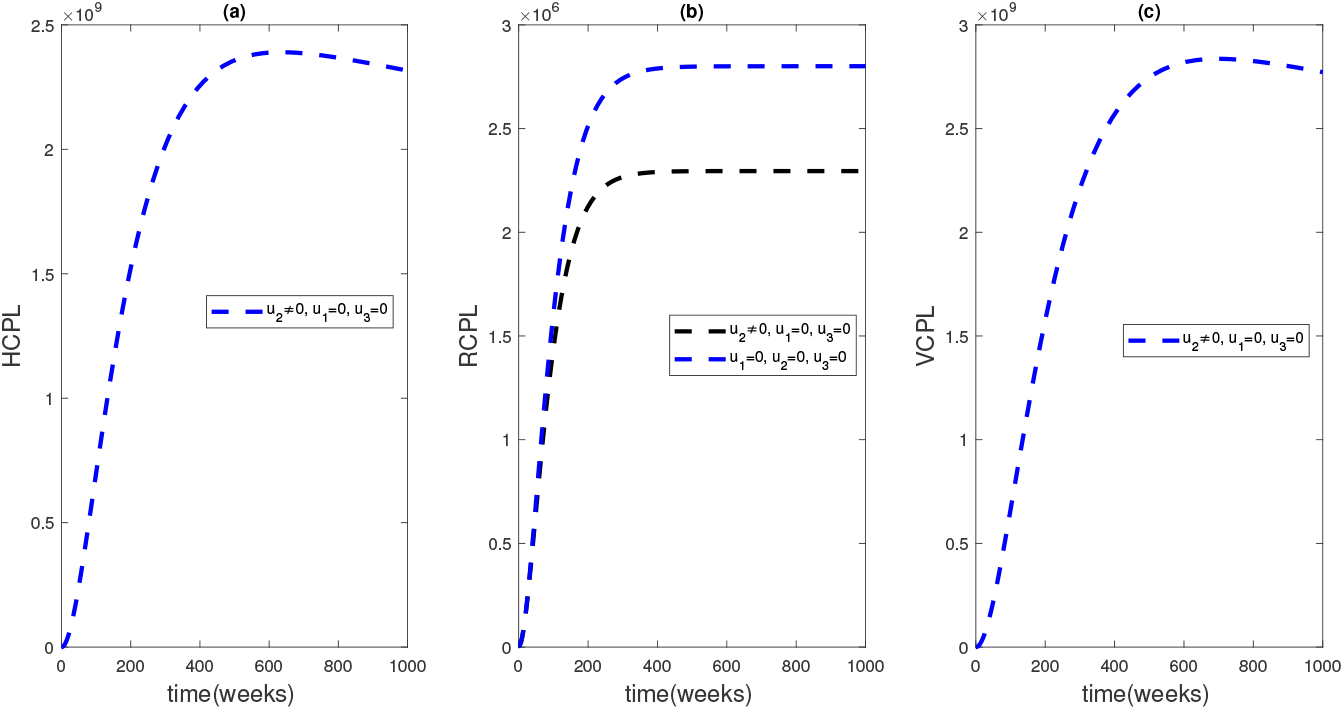
Graphical illustration of the Community Pathogen Load with constant rodent control.

**Fig. 7:**
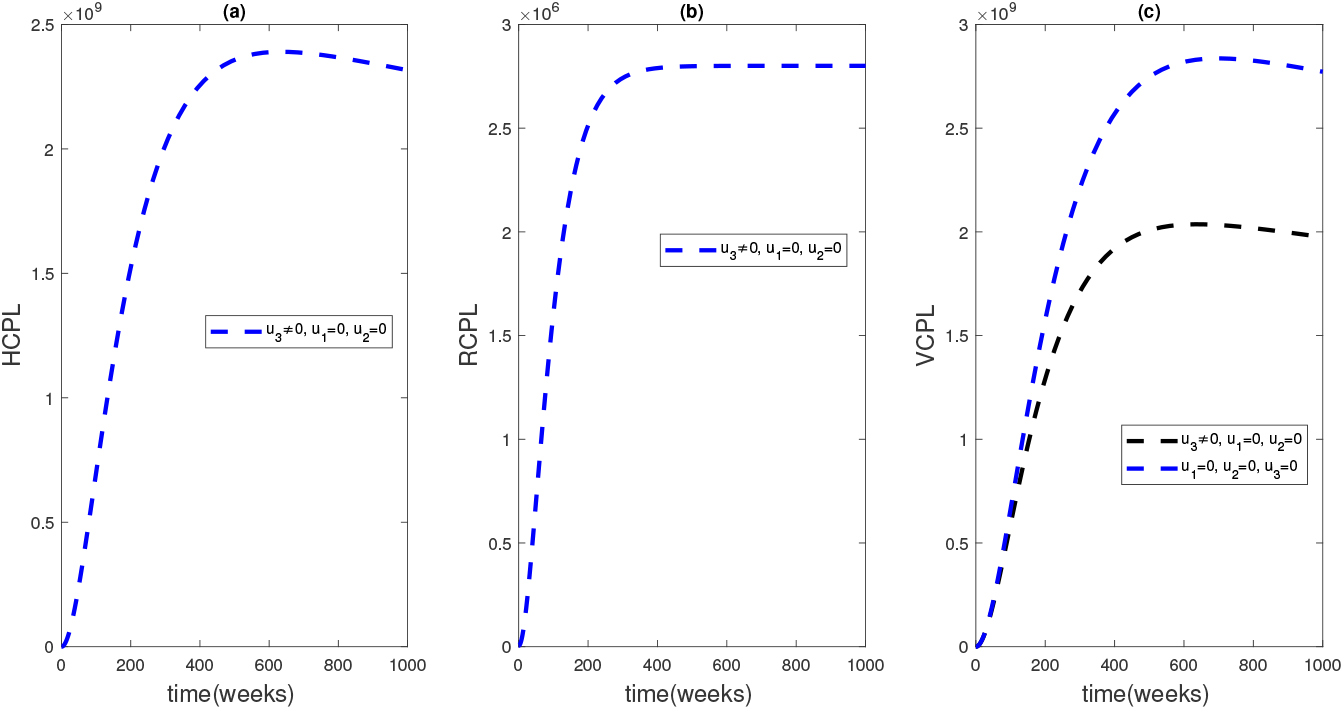
Graphical illustration of the Community Pathogen Load with constant fumigation control.

**Fig. 8:**
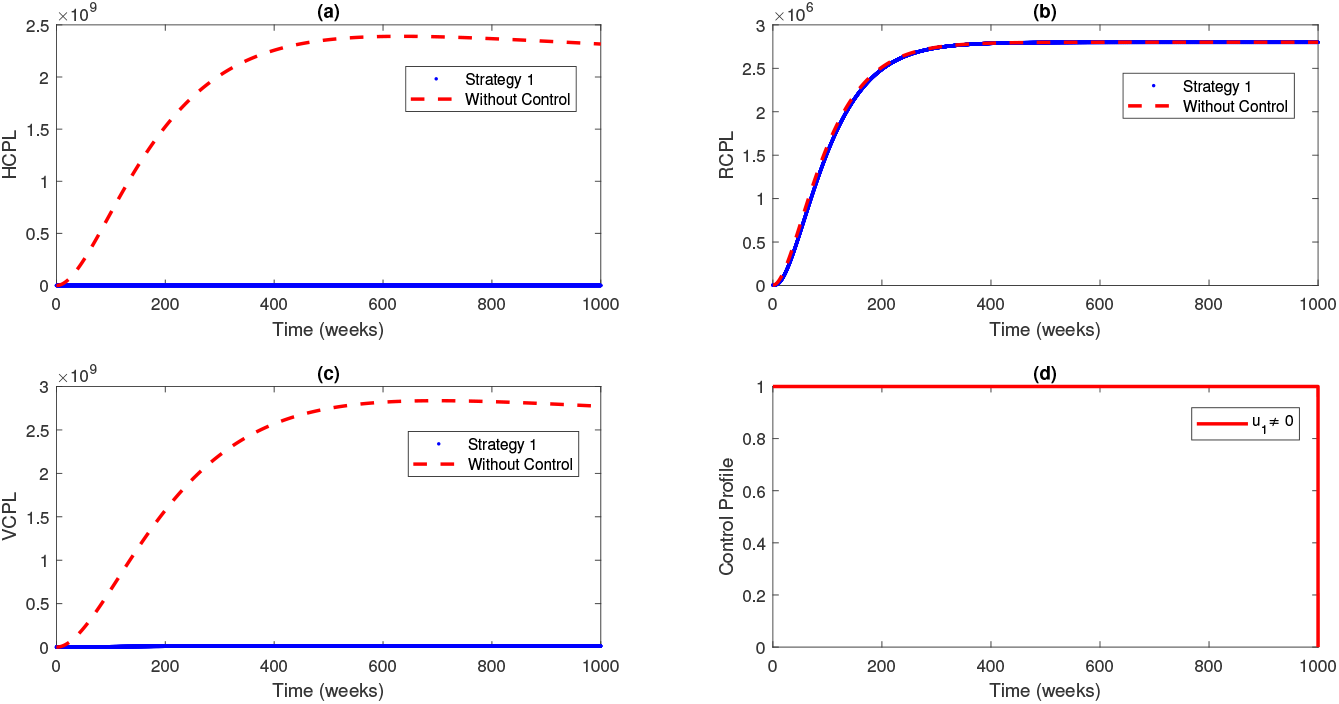
Simulations of the effects of Strategy 1 on HCPL, RCPL, VCPL, and the control profile.

Figure 3 shows the baseline graphs of the system without varying the parameters of the system which is understood as depicting the scenario in which the disease continues to exist in the system. This was done over a time period of 1000 weeks. We see from figure 4 that the introduction of constant control contributes to a decline in the community pathogen load of the human, rodent population and is associated with a little decline in the community pathogen load of the virus population. This, however, cannot be sustained over a long period of time hence the need for optimal control strategies. We observe that the introduction of constant controls is associated with a reduction of the HCPL, RCPL, and VCPL (figures 5 to 7) but the averted infection is smaller compared with the optimal use of time-dependent control strategies (see figures 8 to 10). This result is reasonable since constant control uses fixed control inputs, which may not be efficient at all times and may not adapt to changes in the system dynamics. We illustrate the impacts of the different control strategies in the next subsection.

#### 5.2.1 Numerical Illustration of Control Strategies

In this section, we look at the impact of different control strategies on the objective function by simulating the effect of these strategies on human, rodent, and virus community pathogen load.

(a) Strategy 1: the optimal use of human treatment (with ribavirin), *u*_1_ only. We observe from Figure 8 that human treatment reduces the HCPL and VCPL significantly and has no effect on the RCPL. The control profile of the optimal control *u*_1_ shows that the optimal use of treatment should be maintained at the upper bound 100% throughout the time of intervention.
(b) Strategy 2: the optimal use of rodent control (with mouse traps), *u*_2_ only. Figure 9 shows that rodent control is associated with a decline in the RCPL but has no effect on the HCPL and VCPL. The control profile of the optimal control *u*_2_ shows that the optimal use of rodent control should be sustained throughout the time of intervention with some reduction towards the end of the intervention period.
(c) Strategy 3: the optimal use of fumigation (with pesticides), *u*_3_ only. Figure 10 shows that the use of fumigation is associated with a decrease in the VCPL but has no effect on the HCPL and RCPL. The control profile of the optimal control *u*_3_ shows that the optimal use of fumigation should be sustained at the upper bound 100% throughout the time of intervention.
(d) Strategy 4: the optimal use of human treatment, *u*_1_, and rodent control, *u*_2_, only. Figure 11 shows a decline in the HCPL, RCPL, and VCPL in the presence of the use of treatment and rodent control. The control profile shows that the optimal control *u*_1_ is at its maximum peak throughout the period of intervention. It also shows that the optimal control *u*_2_ is sustained at the upper bound but reduced after some time.
(e) Strategy 5: the optimal use of human treatment, *u*_1_, and fumigation, *u*_3_, only. Figure 12 shows that the use of treatment and fumigation is associated with a decline in the HCPL and VCPL but has no significant effect on the RCPL. The control profile shows that the optimal control *u*_1_ is at its maximum peak throughout the period of intervention. It also shows that the optimal control *u*_3_ rises gradually up to 100% at week 177 and stays at this upper bound till week 2831 when it declines gradually.
(f) Strategy 6: the optimal use of rodent control, *u*_2_, and fumigation, *u*_3_, only. Figure 13 shows a decrease in the RCPL and VCPL when rodent control and fumigation are used but have no effect on HCPL. The control profile shows that the optimal control *u*_2_ is not sustained throughout the time of intervention. Its effect reduces at week 2800 and continues to reduce gradually to about 10%. The optimal control *u*_3_ rises gradually up to 100% at the beginning of the intervention and stays at this upper bound throughout the period of intervention.
(g) Strategy 7: the optimal use of human treatment, *u*_1_, rodent control, *u*_2_, and fumigation, *u*_3_, only. Figure 14 shows a decline in the HCPL, RCPL, and VCPL using a combination of treatment, rodent control, and fumigation. The control profile shows that the optimal control *u*_1_ remains at the upper bound throughout the time of intervention. The effect of *u*_2_ starts at the upper bound from the beginning of intervention but begins to decline from week 2377 till about 5%. The optimal control *u*_3_ rises slowly from 4% at the beginning of the intervention, settles at the upper bound, and then begins to wean at week 1508 till it reaches the lower bound at the end of the period of intervention.

**Fig. 9:**
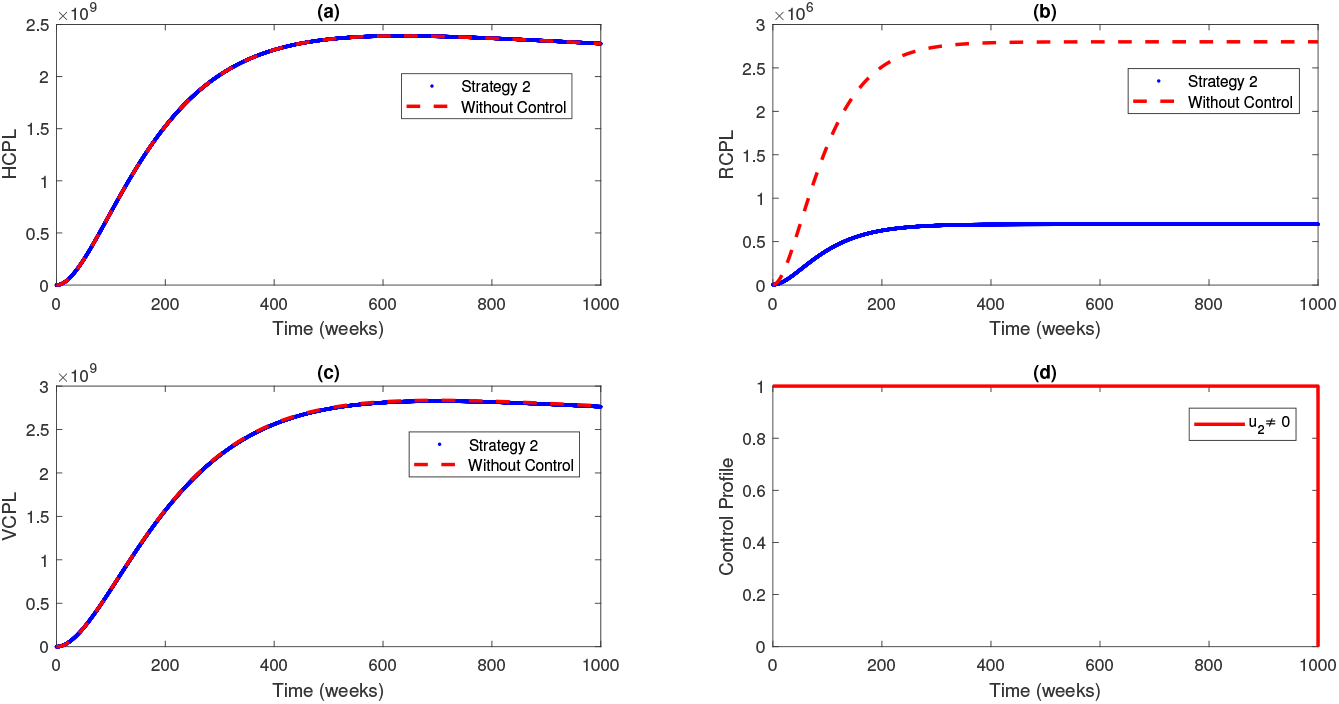
Simulations of the effects of Strategy 2 on HCPL, RCPL, VCPL, and the control profile.

**Fig. 10:**
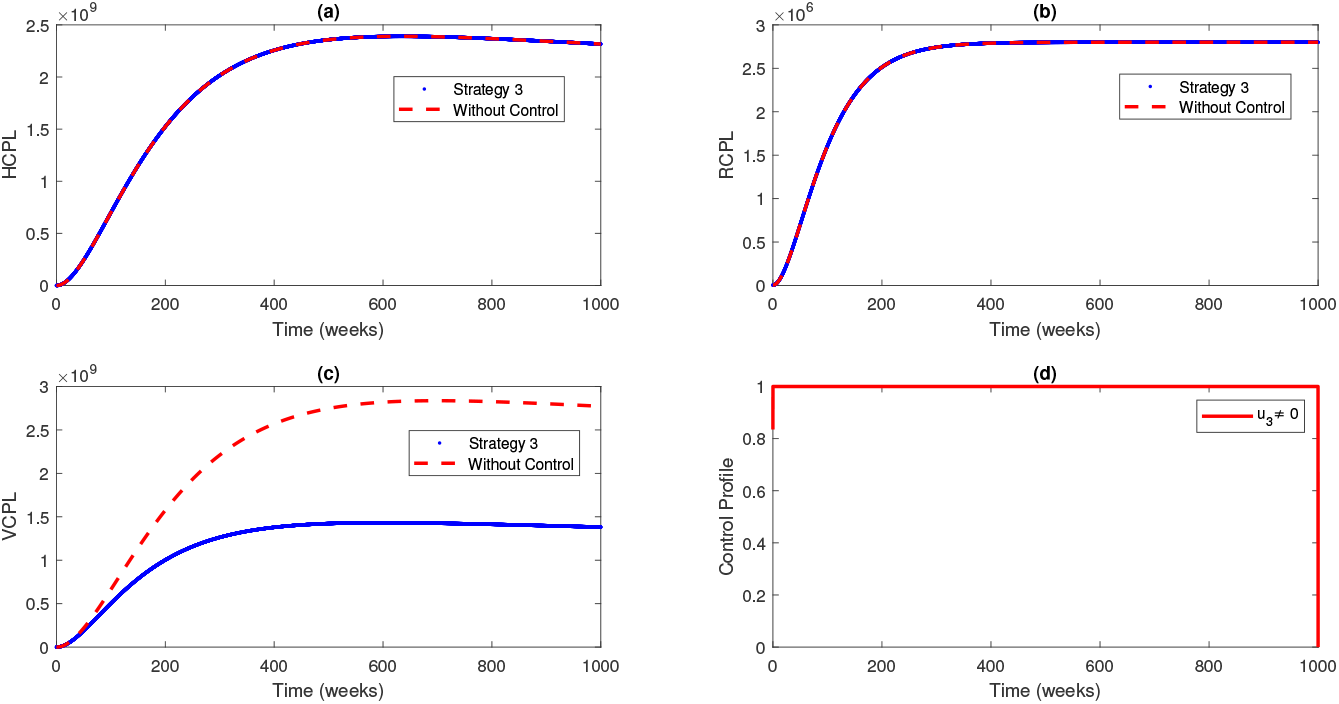
Simulations of the effects of Strategy 3 on HCPL, RCPL, VCPL, and the control profile.

**Fig. 11:**
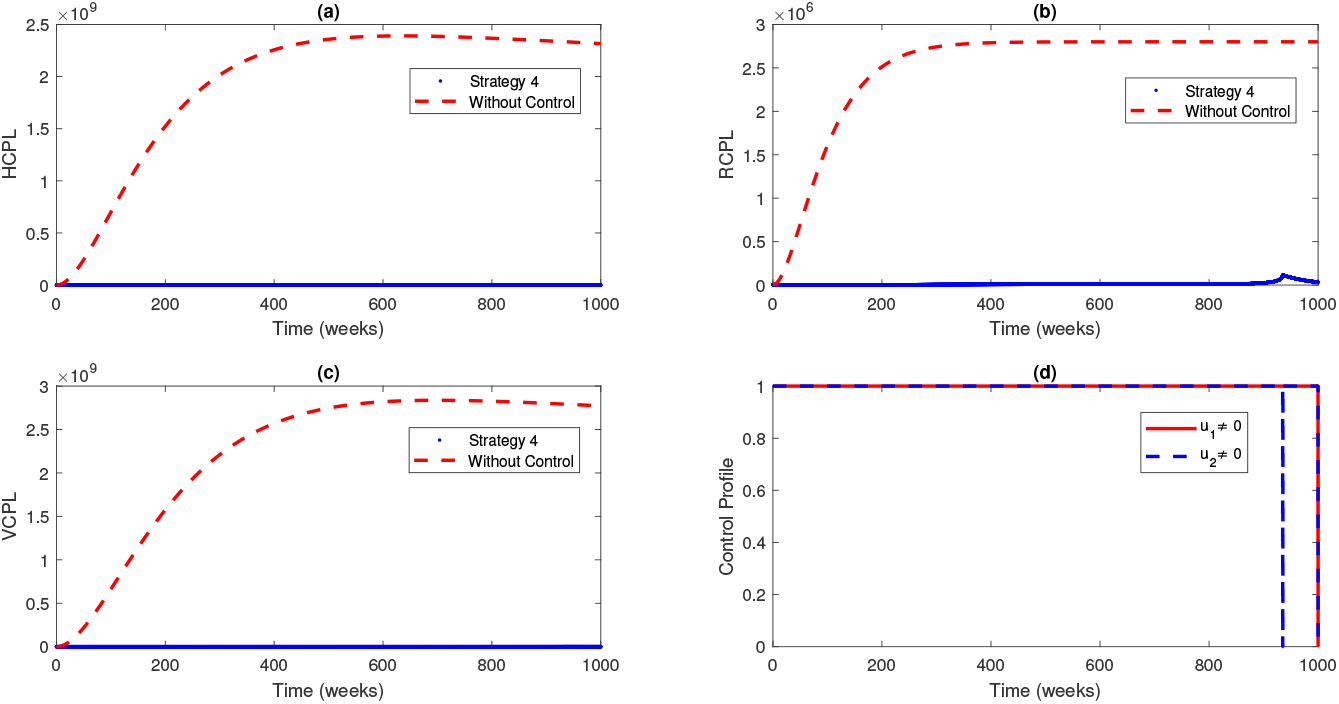
Simulations of the effects of Strategy 4 on HCPL, RCPL, VCPL, and the control profile.

**Fig. 12:**
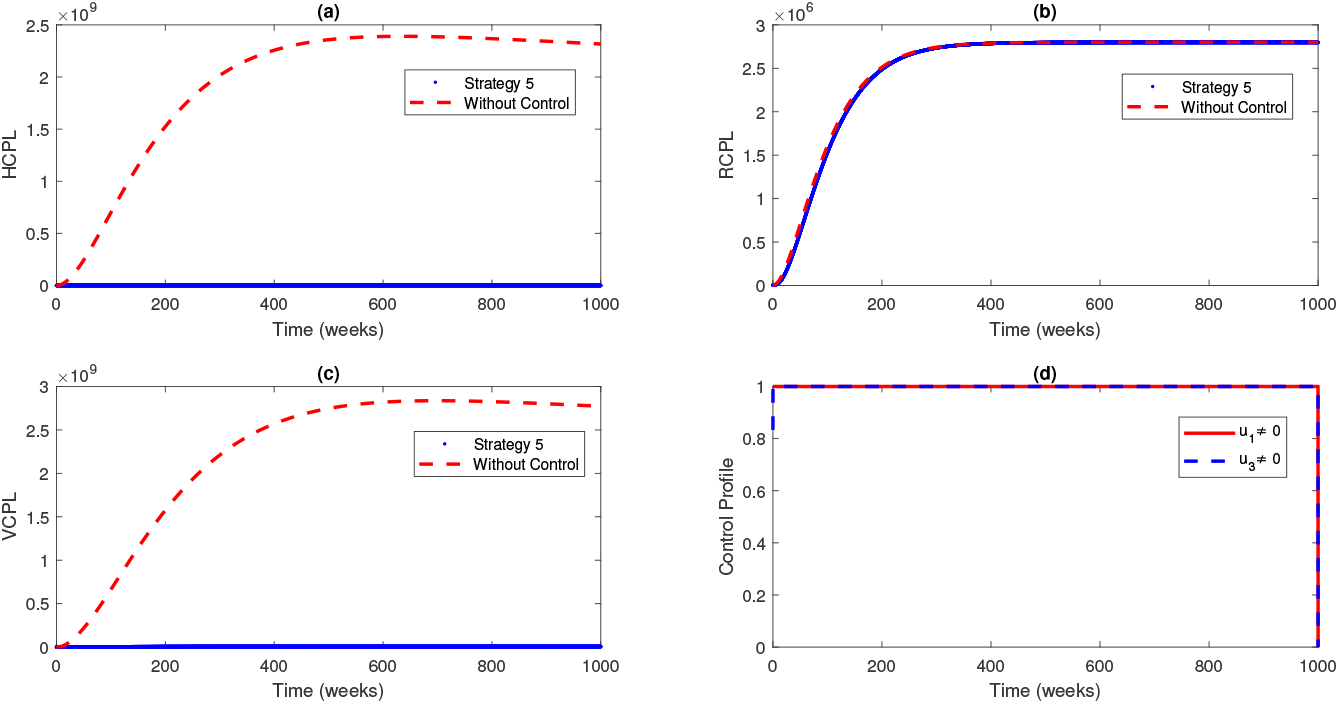
Simulations of the effects of Strategy 5 on HCPL, RCPL, VCPL, and the control profile.

**Fig. 13:**
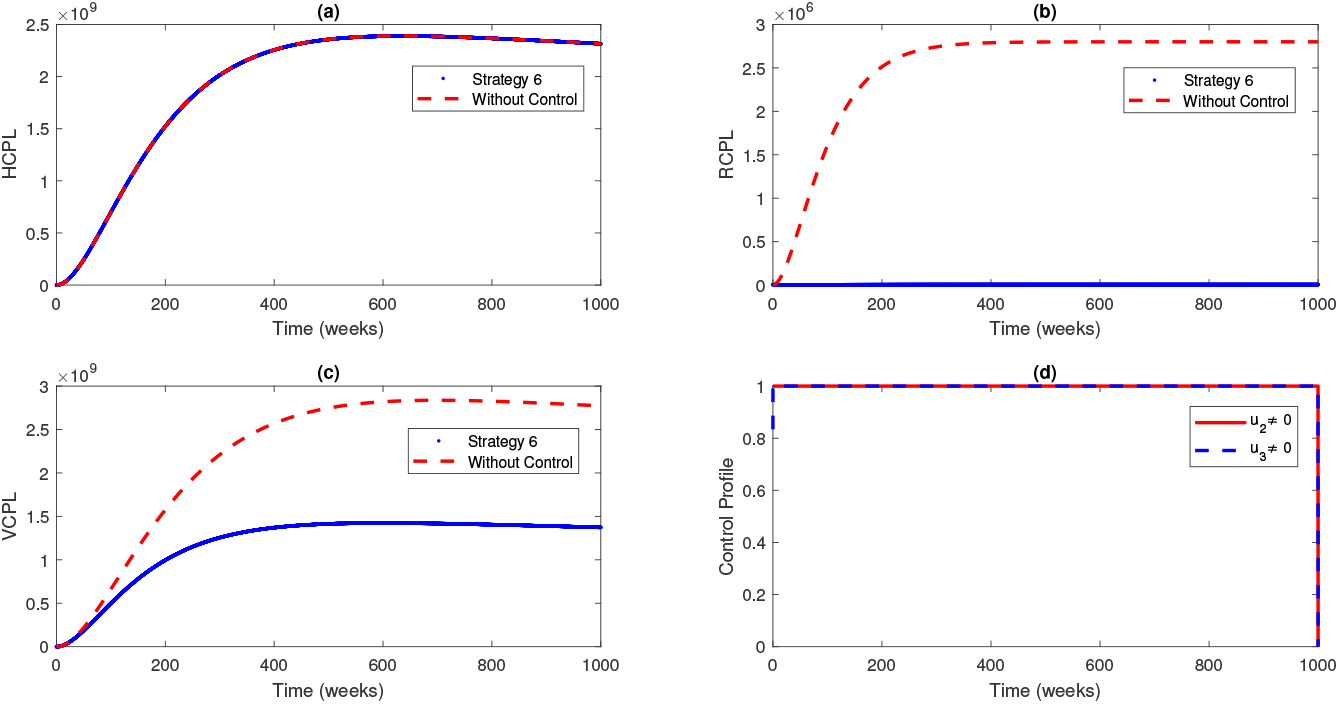
Simulations of the effects of Strategy 6 on HCPL, RCPL, VCPL, and the control profile.

**Fig. 14:**
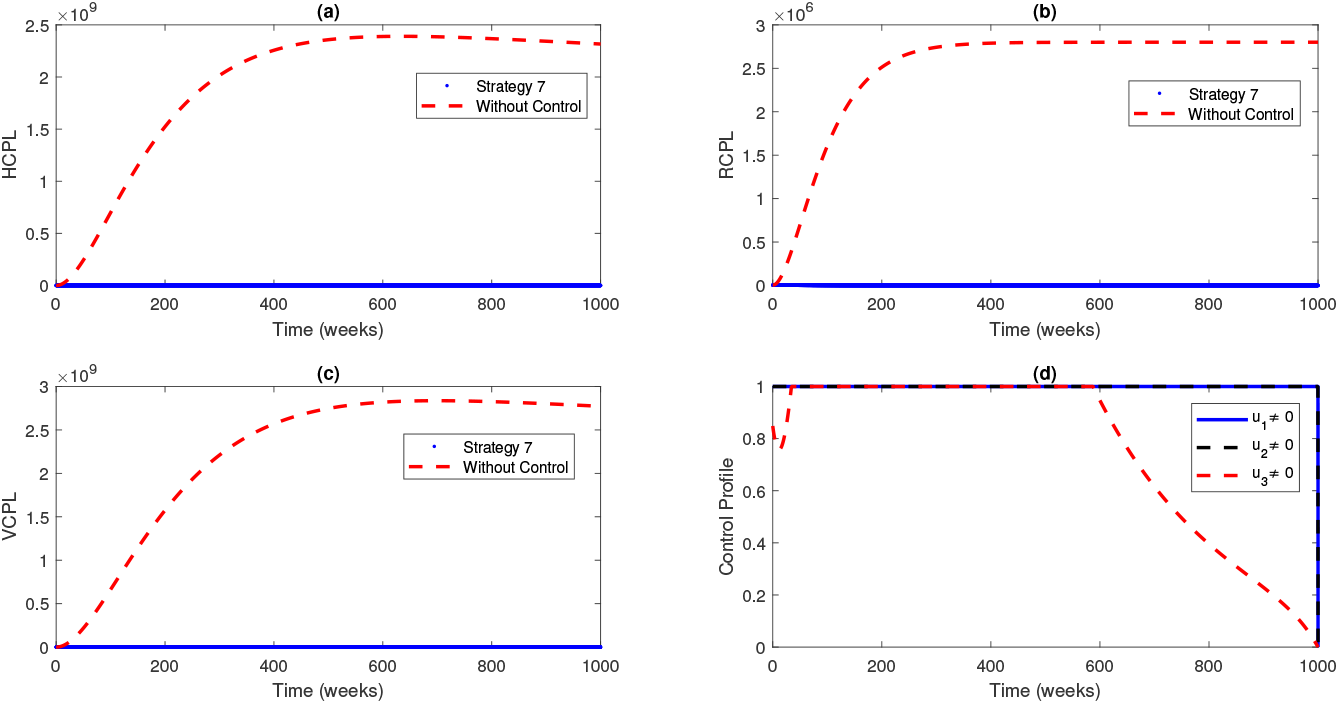
Simulations of the effects of Strategy 7 on HCPL, RCPL, VCPL, and the control profile.

We observe that the effect of different control strategies (single, double, triple) reduces the community pathogen load in the human, rodent, and virus populations. It is important to implement at least a single control strategy in the population. It is noteworthy to mention that some single-control strategies have a greater effect than others in terms of averting infection. For instance, the use of human treatment affects both HCPL and VCPL. In areas with limited resources, this suggests prioritizing treatment to fumigation. The use of rodent control also averts greater infection than the use of fumigation only. So, by ranking the single controls, we see that human treatment has the greatest effect, followed by rodent control and then fumigation. For the double controls, strategy 4 has the greatest effect, followed by strategy 5 and then strategy 6. Considering the infection averted by implementing each strategy alongside the cost involved, we look at the best cost-effective strategy for this in the next section.

### 5.3 Cost Effective Analysis

In developing countries like Nigeria, resources for funding control interventions can be scarce and expensive. Healthcare interventions should be assessed by comparing the costs and outcomes associated with different strategies. The Cost Effective Analysis (CEA) is useful in making informed decisions about allocating limited resources to interventions that are likely to provide the most value in terms of improving health outcomes. In this section, we use different approaches like infection averted ratio (IAR), average cost-effectiveness ratio (ACER), and incremental cost-effectiveness ratio (ICER) [42, 43] to achieve this aim.

#### 5.3.1 Infection Averted Ratio (IAR)

IAR is the ratio of the number of infections averted by an intervention to the number of people who recovered from the infection as a result of the control strategies.

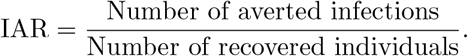

The difference between the total number of infected individuals without optimal control and the total number of infectious individuals with optimal control is referred to as the number of infections averted. Strategies with the highest IAR are considered as the most cost-effective in reducing the spread of the disease within the population [42]. From table 3, the strategies that are cost-effective are strategies 1, 4, 5, and 7.

**Table 3:**
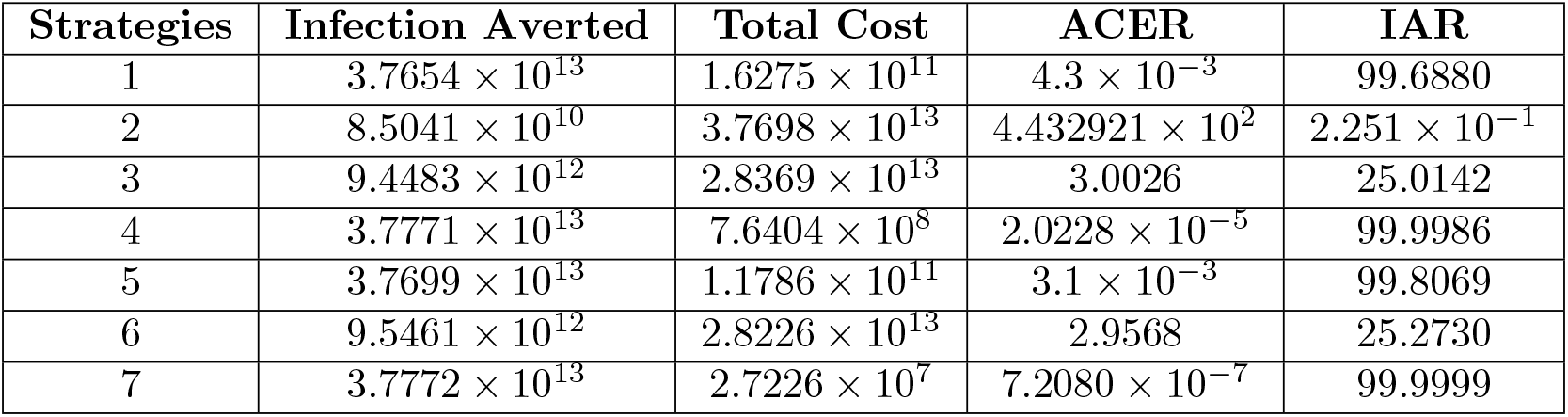
Total cost, total infection averted, IAR, and ACER of all interventions.

#### 5.3.2 Average Cost Effectiveness Ratio (ACER)

ACER is the ratio of the total cost of implementing a particular intervention to the total number of infections averted by that intervention. So,

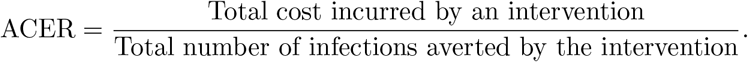

Interventions with the lowest ACER scores are the most cost-effective and strategies with the highest ACER scores are the least cost-effective. Table 3 shows that strategies 1, 4, 5, and 7 are the most cost-effective interventions.

#### 5.3.3 Incremental Cost Effectiveness Ratio (ICER)

ICER is used to determine the difference between two or more competing strategies, along with health benefits and implementation costs. It helps to obtain the intervention with the minimum cost across all available schemes given in this study. It is given by

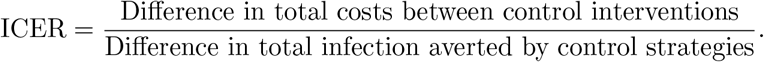

We investigate the most cost-effective strategy among the single, double, and triple interventions. The single interventions are strategy 1, strategy 2, and strategy 3. The ICER calculations are as follows:

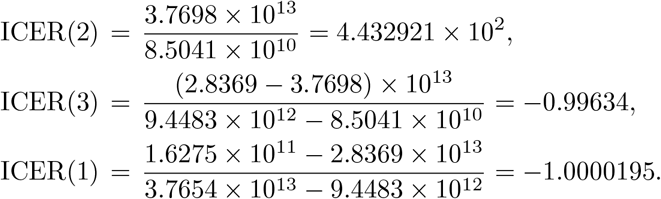

Comparing strategies 2 and 3, the ICER(2) is greater than ICER(3) showing that strategy 2 is more expensive to implement. We eliminate strategy 2 and continue our investigation with the remaining strategies as follows:

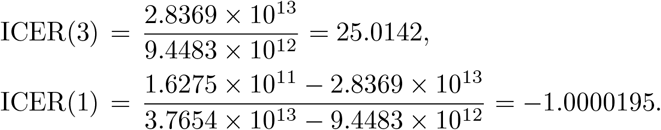

Since ICER(3) is bigger than ICER(1), it shows that strategy 1 is less costly than strategy 3. Among the single interventions, we see that strategy 1 is the most cost-effective strategy. This is in agreement with the IAR and ACER method used.

The double interventions include strategy 4, strategy 5, and strategy 6. We perform the ICER calculations as follows:

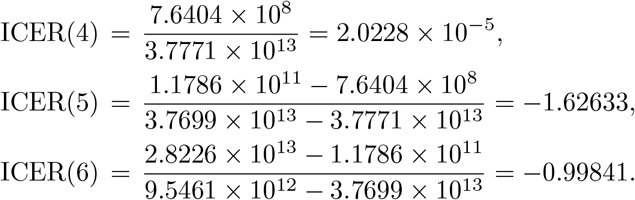

When we compare strategies 4 and 5, we see that ICER(4) is bigger than ICER(5). This shows that strategy 4 is more expensive to execute. We remove strategy 4 and continue with the remaining strategies as follows:

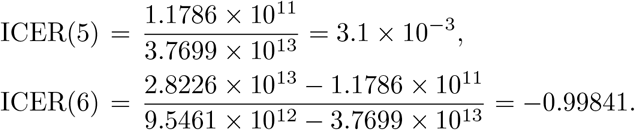

Strategy 6 is less expensive than strategy 5 since ICER(5) *>* ICER(6). Hence, among the double interventions, strategy 6 is the most cost-effective strategy. This agrees with the IAR and ACER methods used.

In order to get the minimum cost strategy across all schemes, we have to perform the ICER calculations among the outstanding schemes such as strategy 1, strategy 6, and strategy 7. We present our results in table 4. From table 4, we see that ICER(1) *>* ICER(6) which shows that strategy 1 is more expensive and less effective to execute compared to strategy 6. We remove Strategy 1 and continue our investigation with the remaining strategies. These ICER results are shown in table 5. The result from table 5 shows that strategy 6 dominates strategy 7 so it is more expensive to implement. There, the multiple intervention (strategy 7) is the most cost-effective among all the strategies.

**Table 4:**
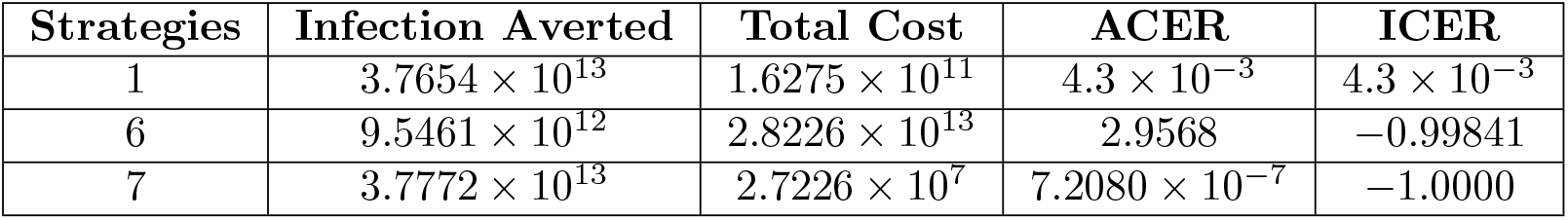
ICER for strategies 1, 6, and 7.

**Table 5:**
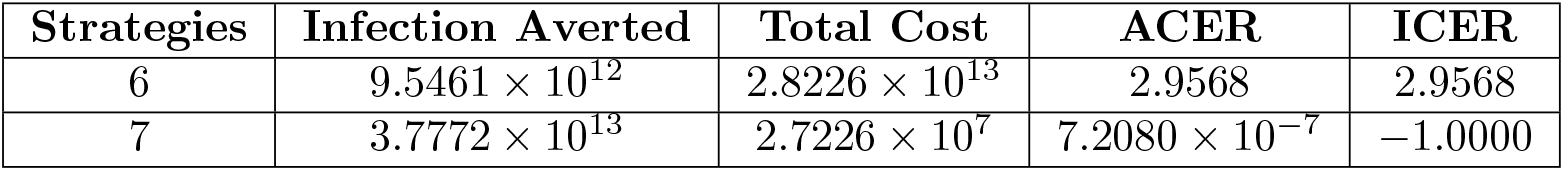
ICER for strategies 6 and 7.

| Strategies | Infection Averted | Total Cost | ACER | ICER |
| --- | --- | --- | --- | --- |
| 6 | $9.5461 \times 10^{12}$ | $2.8226 \times 10^{13}$ | 2.9568 | 2.9568 |
| 7 | $3.7772 \times 10^{13}$ | $2.7226 \times 10^7$ | $7.2080 \times 10^{-7}$ | -1.0000 |

### 5.4 Sensitivity Analysis of Parameters on Control Profile

We perform a local sensitivity analysis on the control profile. We investigate the impact of a few parameters on the trajectory of the optimal control profile of the single control strategies.

As we investigate the parameter changes on the control profiles, we also present the averted infection shown by these changes in table 6. We perturbed input parameters to observe the resulting changes in the control profiles. Most of the parameters we perturbed did not alter the behavior of the control profiles so we only presented the outputs of the control profiles where perturbed parameters produced changes in the trajectories.

**Table 6:** Averted Infection for parameter variations.

| Strategies | Parameter | Infection Averted |
| --- | --- | --- |
| 1 | $\tau_H = 0.3$ | $9.0222 \times 10^3$ |
| | $\tau_H = 0.003$ | $9.0222 \times 10^3$ |
| | $\tau_H = 0.00003$ | $9.0215 \times 10^3$ |
| 2 | $\alpha_R = 0.3086$ | $8.2343 \times 10^3$ |
| | $\alpha_R = 0.03086$ | $8.2341 \times 10^3$ |
| | $\alpha_R = 0.003086$ | $8.2340 \times 10^3$ |
| 2 | $\tau_R = 0.1005$ | $8.4234 \times 10^3$ |
| | $\tau_R = 0.01005$ | $8.4232 \times 10^3$ |
| | $\tau_R = 0.001005$ | $8.4231 \times 10^3$ |

From figure 15, we see that with a smaller value of *τ*_*H*_, the control profile of *u*_1_ operates at 10% at the beginning of the intervention. With a lower production rate of *V*_*H*_ virus in one infected person, we need not invest so much in strategy 1 since the infection averted will also be lower according to table 6. Figure 16 shows that the growth rate of Lassa Fever viral load in infected rodents and the production rate of *V*_*R*_ virus in one infected rodent is associated with the trajectory of strategy 2. When these rates are low, our intervention will not operate at 100% capacity so we do not need to invest so much in them since the effect of the intervention will not be sustained over a long period of time. Generally, table 6 shows us the different changes in averted infection with respect to parameter variations. The averted infection is not only dependent on the control strategies introduced but also on variations in the underlying parameters used.

**Fig. 15:**
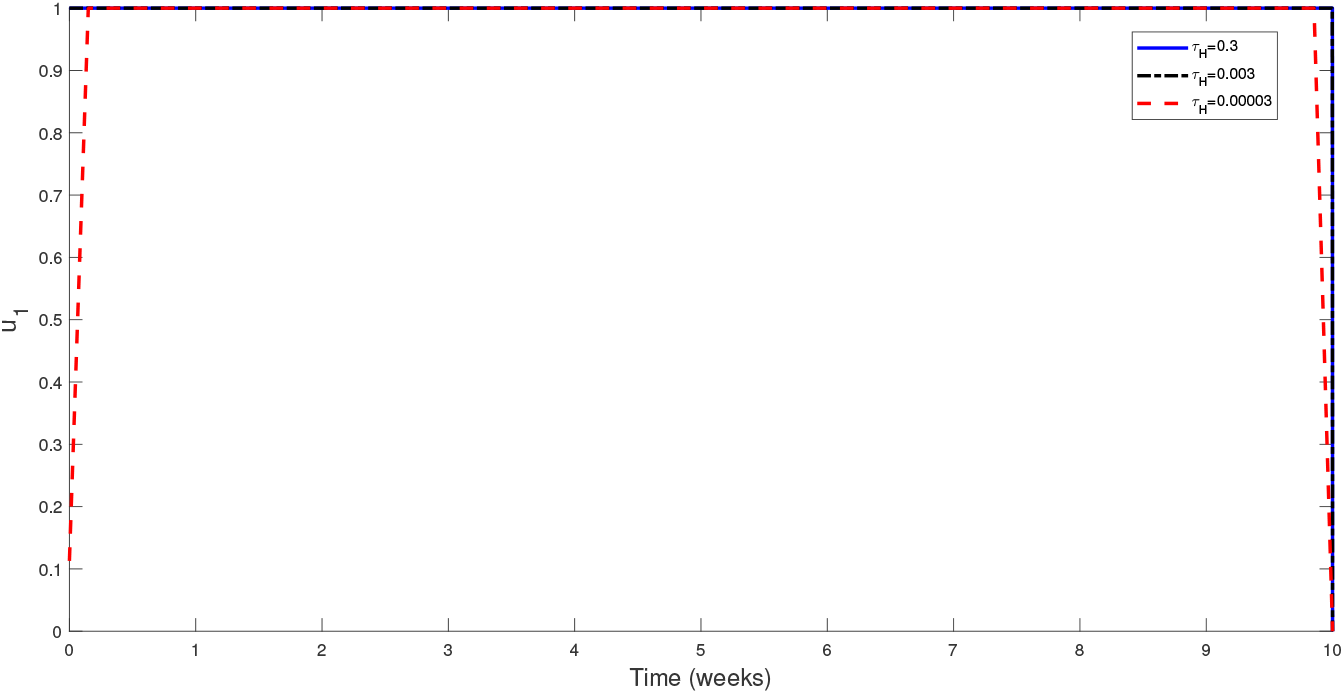
The effect of τ_*H*_ on the control profile *u*_1_.

**Fig. 16:**
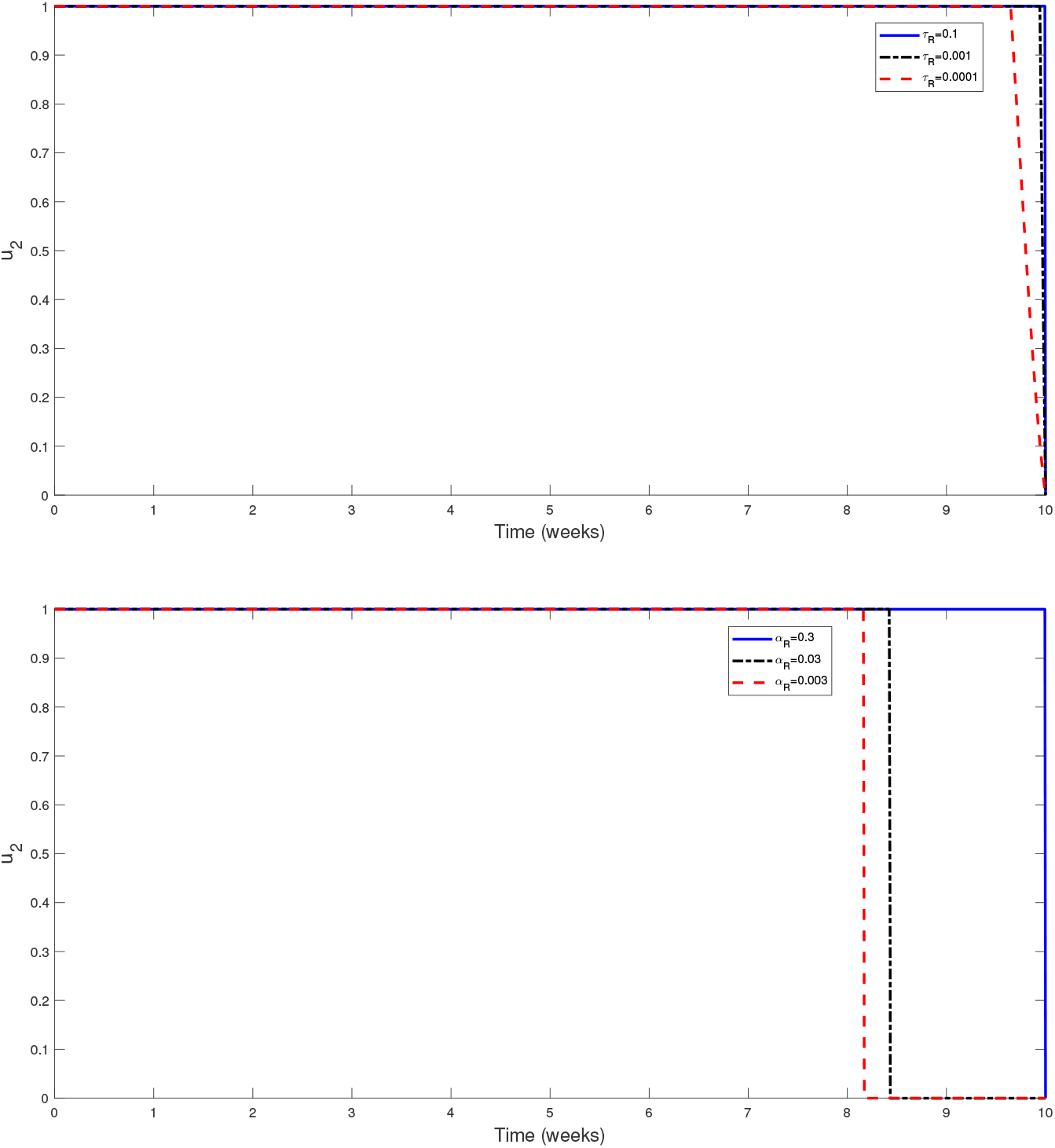
The effect of *τ*_*R*_ and *α*_*R*_ on the control profile *u*_2_.

## 6 Discussion of Results

In this study, we introduced a community pathogen load (CPL) method to the transmission dynamics of Lassa Fever infection. This method is based on assumptions about how individual infectivity is scaled up to define population/community infectivity. Therefore, the model developed using this method not only tracks the number of infected hosts but also considers the actual parasite load at the host-to-host level. We used mathematical tools to establish the local stability of the endemic steady state and the global stability of the disease-free steady state. Our analysis yielded mathematical results that indicate the conditions under which disease persists or can be controlled within the system and the sensitivity to parameter changes in the optimal control profile in the course of system dynamics. The control strategies introduced were targeted at the reduction of the community contribution of Lassa viral load from the human, rodent, and environmental virus populations. While the use of constant controls reduces the pathogen load in all populations, we observed that it is not an adequate measure of control as it does not account for adjusted controls due to variations in infection dynamics over time. Results from our simulations showed that the optimal use of human treatment should be given priority, especially in communities with limited resources. Our study also showed that the multiple intervention strategy within controls is the best measure for controlling the spread of Lassa Fever and also the most cost-effective since it averts a greater quantity of infection with minimal cost. This is significant since the community pathogen load contribution from human, rodent, and virus populations cannot be curbed using just one control measure. In developing countries like Nigeria, this might not be easy considering it has a vast number of rural areas with limited resources. However, we recommend that public health policies should be targeted at distributing resources appropriately so that every intervention strategy will be accounted for as seen from our Cost-effective and optimal control analysis. Our results support the use of community pathogen load as a metric that can be used for Lassa fever control targets in both humans and rodents as well as the viral load in the environment. The combination of the three measures (IAR, ACER, ICER) in our Cost-Effective Analysis provided a more complete picture and a comprehensive analysis of the cost-effectiveness of different control strategies as tools that can be used to evaluate the benefits of various control strategies viz a viz the costs of implementation. The three metrics are important in estimating different aspects of disease control in health economics and disease management. The infection averted ratio is essential in measuring the effectiveness of interventions in preventing infections and important in outbreak response. The average cost-effectiveness ratio is important in evaluating the effectiveness of intervention in situations with no clear comparisons between new and existing/alternative control measures. The incremental cost-effectiveness ratio is a metric that is useful in evaluating the benefits of using one intervention compared to another in situations where there is a clear comparison criteria between new and existing/alternative control measures. In our study, the three metrics were in agreement in predicting the combination of three control strategies as the best cost-effective strategy in averting Lassa Fever infection.

The current study was just a precursor to the multiscale modeling of Lassa Fever infection in human-rodent transmission with environmental components. There is a need to incorporate the within-host stages of infection to account for stage-specific and scale-specific transmission impact as well as intervention strategies. This can be coupled with various aspects that affect disease prognosis such as climate change.

## Data Availability

All data used are available online at https://www.ncdc.gov.ng/diseases/sitreps/.

## Declaration of Interest

The authors declare that there is no conflict of interest regarding the submitted manuscript. All research was done independently and no external funding or competing interests influenced the design, analysis, write-up, and results discussion of the manuscript.

## Supplementary information

Not applicable.

## Acknowledgements

Praise-God was supported by the University of Johannesburg’s URC funds. F. Chirove was supported by the NRF IPRR grant with grant N0: 132253.

## Declarations

## Appendix A Section title of first appendix

Not applicable.

## Notes

### Competing Interest Statement

The authors have declared no competing interest.

### Funding Statement

This study was funded by University of Johannesburg URC funds and NRF IPRR grant with grant number, 132253

